# Integrative Analysis of Germline Rare Variants in Clear and Non-Clear Cell Renal Cell Carcinoma

**DOI:** 10.1101/2023.01.18.23284664

**Authors:** Seunghun Han, Sabrina Y. Camp, Hoyin Chu, Ryan Collins, Riaz Gillani, Jihye Park, Ziad Bakouny, Cora A. Ricker, Brendan Reardon, Nicholas Moore, Eric Kofman, Chris Labaki, David Braun, Toni K. Choueiri, Saud H. AlDubayan, Eliezer M. Van Allen

## Abstract

**IMPORTANCE:** RCC encompasses a set of histologically distinct cancers with a high estimated genetic heritability, of which only a portion is currently explained. Previous rare germline variant studies in RCC have usually pooled clear and non-clear cell RCCs and have not adequately accounted for population stratification that may significantly impact the interpretation and discovery of certain candidate risk genes.

**OBJECTIVE:** To evaluate the enrichment of germline PVs in established cancer-predisposing genes (CPGs) in clear cell and non-clear cell RCC patients compared to cancer-free controls using approaches that account for population stratification and to identify unconventional types of germline RCC risk variants that confer an increased risk of developing RCC.

**DESIGN, SETTING, AND PARTICIPANTS:** In 1,436 unselected RCC patients with sufficient data quality, we systematically identified rare germline PVs, cryptic splice variants, and copy number variants (CNVs). From this unselected cohort, 1,356 patients were ancestry-matched with 16,512 cancer-free controls, and gene-level enrichment of rare germline PVs were assessed in 143 CPGs, followed by an investigation of somatic events in matching tumor samples.

**MAIN OUTCOMES AND MEASURES:** Gene-level burden of rare germline PVs, identification of secondary somatic events accompanying the germline PVs, and characterization of less-explored types of rare germline PVs in RCC patients.

**RESULTS:** In clear cell RCC (n = 976 patients), patients exhibited significantly higher prevalence of PVs in *VHL* compared to controls (OR: 39.1, 95% CI: 7.01-218.07, p-value:4.95e-05, q-value:0.00584). In non-clear cell RCC (n = 380 patients), patients carried enriched burden of PVs in *FH* (OR: 77.9, 95% CI: 18.68-324.97, p-value:1.55e-08, q-value: 1.83e-06) and *MET* (OR: 1.98e11, 95% CI: 0-inf, p-value: 2.07e-05, q-value: 3.50e-07). In a *CHEK2*-focused analysis with European cases and controls, clear cell RCC patients (n=906 European patients) harbored nominal enrichment of the previously reported low-penetrance *CHEK2* variants, p.Ile157Thr (OR:1.84, 95% CI: 1.00-3.36, p-value:0.049) and p.Ser428Phe (OR:5.20, 95% CI: 1.00-26.40, p-value:0.045) while non-clear cell RCC patients (n=295 European patients) exhibited nominal enrichment of *CHEK2* LOF germline PVs (OR: 3.51, 95% CI: 1.10-11.10, p-value: 0.033). RCC patients with germline PVs in *FH, MET, and VHL* exhibited significantly earlier age of cancer onset compared to patients without any germline PVs in CPGs (Mean: 46.0 vs 60.2 years old, Tukey adjusted p-value < 0.0001), and more than half had secondary somatic events affecting the same gene (n=10/15, 66.7%, 95% CI: 38.7-87.0%). Conversely, patients with rare germline PVs in *CHEK2* exhibited a similar age of disease onset to patients without any identified germline PVs in CPGs (Mean: 60.1 vs 60.2 years old, Tukey adjusted p-value: 0.99), and only 30.4% of the patients carried secondary somatic events in *CHEK2* (n=7/23, 95% CI: 14.1-53.0%). Finally, rare pathogenic germline cryptic splice variants underexplored in RCC were identified in *SDHA* and *TSC1*, and rare pathogenic germline CNVs were found in 18 patients, including CNVs in *FH, SDHA*, and *VHL*.

**CONCLUSIONS AND RELEVANCE:** This systematic analysis supports the existing link between several RCC risk genes and elevated RCC risk manifesting in earlier age of RCC onset. Our analysis calls for caution when assessing the role of germline PVs in *CHEK2* due to the burden of founder variants with varying population frequency in different ancestry groups. It also broadens the definition of the RCC germline landscape of pathogenicity to incorporate previously understudied types of germline variants, such as cryptic splice variants and CNVs.

**KEY Points:** 

**Question:** Can we improve the assessment of germline genetic risk determinants for clear cell and non-clear cell renal cell carcinoma (RCC) with approaches that are aware to population-stratification and RCC histological subtypes?

**Findings:** In this systematic case-control study of 1,356 RCC patients and 16,512 ancestry-matched cancer-free controls strictly controlling for population stratification, clear-cell RCC patients exhibited a significantly higher prevalence of rare germline pathogenic variants (PVs) in *VHL*, and non-clear cell RCC patients carried significantly more rare germline PVs in *FH* and *MET*. European clear-cell RCC patients harbored a nominally significant enrichment of two low-penetrance *CHEK2* variants (p.Ser428Phe and p.Ile157Thr) while European non-clear RCC patients carried a nominally significant enrichment of rare germline loss-of-function (LOF) variants in *CHEK2*. Subsequent somatic analyses identified secondary somatic events in genes significantly enriched for germline PVs (*VHL, FH, MET*), and these variant carriers presented with earlier age of disease onset, but *CHEK2* germline variant carriers harbored relatively fewer somatic events in *CHEK2* and did not present with earlier age of onset. Finally, we identified 6 RCC patients with rare germline cryptic splice and copy number variants that impacted known kidney cancer risk genes, increasing the diagnostic yield of pathogenic variants in RCC risk genes from 2.1% to 2.5%.

**Meaning:** Clear and non-clear RCCs have distinct germline pathogenic variant enrichment patterns and somatic variants. Accurate risk assessment of *CHEK2* in RCC requires careful adjustment for population stratification. In addition, previously underappreciated forms of germline variants may explain a portion of the missing heritability in RCC.

## INTRODUCTION

Renal cell carcinoma (RCC) is the ninth most common neoplasm in the United States, accounting for 2% of all cancers worldwide^1^. The Nordic Twin study has placed the genetic heritability of RCC as high as 38%^2^, however, only a fraction of the heritability is explained by currently identified rare and common RCC risk loci. Moving beyond known RCC risk genes (eTable 1)^3,4^, several pan-RCC studies have reported rare germline PVs in DNA damage repair (DDR) genes such as *CHEK2, ATM*, or *BRCA1/2* ^5-10^, suggesting that inherited defects in DDR may contribute to RCC risk. However, most studies lacked ancestry-matched cancer-free controls to formally test these hypotheses.

Two recent studies performed case-control gene-level burden analyses, in RCC alone^11^ and across cancer types, finding a higher burden of germline PVs in *CHEK2* in RCC patients compared to matched controls^12^. However, these studies pooled all RCC subtypes together as one phenotype for association testing, although clear cell RCC (ccRCC) and non-clear cell RCCs (nccRCC, e.g., papillary, chromophobe) have distinct molecular and clinical features^13,14^. Furthermore, additional analyses are necessary to account for fine-level population stratification within Europe to mitigate spurious association^15^, especially when evaluating genes like *CHEK2*, which is known to harbor many putative PVs that are founder variants from bottlenecked populations (e.g., Ashkenazi Jewish) with highly variable allele frequencies between different European sub-populations.

Here, we first performed a germline variant discovery analysis of 1,436 unselected RCC patients to characterize several types of genomic variation. Next, we performed a case-control association study of ccRCC and nccRCC in a subset (n=1,356) that was successfully ancestry-matched with 16,512 cancer-free controls, and we utilized an ancestry-informed generalized linear model (GLM) to evaluate the major germline drivers of RCC risk. Furthermore, we performed a sub-European ancestry-focused meta-analysis of *CHEK2* to address finer-level population stratification within European populations, and we evaluated associated tumor genomic data for concomitant somatic assessments of candidate PVs. Finally, we evaluated potential clinically relevant but underexplored germline variant types (cryptic splice and copy number variants) by using integrative genomic and transcriptomic analyses, all toward expanding and refining the landscape of germline pathogenic variation in RCC.

## METHODS

### RCC Case Cohort

Whole-exome sequencing (WES) Binary Alignment Maps (BAMs) aligned to Genome Research Consortium human build 37 (GRCh37) from 1,436 RCC patients were collected from 8 different RCC studies (Figure 1; Table 1). Samples from TCGA-KIRC^16^, TCGA-KIRP^17^, and TCGA-KICH^18^ studies were analyzed using the germline BAMs stored in the controlled access TCGA workspaces in the Terra analysis platform (https://terra.bio/). Germline BAMs from CheckMate-025^19^ (NCT01668784, EGAD00001006029), CheckMate-010^20^ (NCT01354431, EGAS00001004291), CheckMate-009^21^ (NCT01358721, EGAD00001006027) were downloaded from European Genome-Phenome Archive (EGA) data repository. Whole genome sequencing (WGS) BAMs of ccRCC patients were downloaded from the International Network of Cancer Genome Consortium (ICGC) PanCancer Analysis of Whole Genomes (PCAWG) Dataset^22^: Renal Cell Cancer – EU/FR (RECA-EU) PCAWG WGS (Dataset ID: EGAD00001002131) and were sliced using a custom exome-target intervals to only include coding regions captured in the WES samples. Data from papillary and chromophobe RCC patients from Genentech were downloaded from EGA (Dataset ID: EGAD00001001023^23^). All samples underwent identical quality control procedures and were processed using the same analytical methods. This study was approved by the participating institutions where written consent from participants was collected. This study conforms to the Declaration of Helsinki.

**Figure 1.**
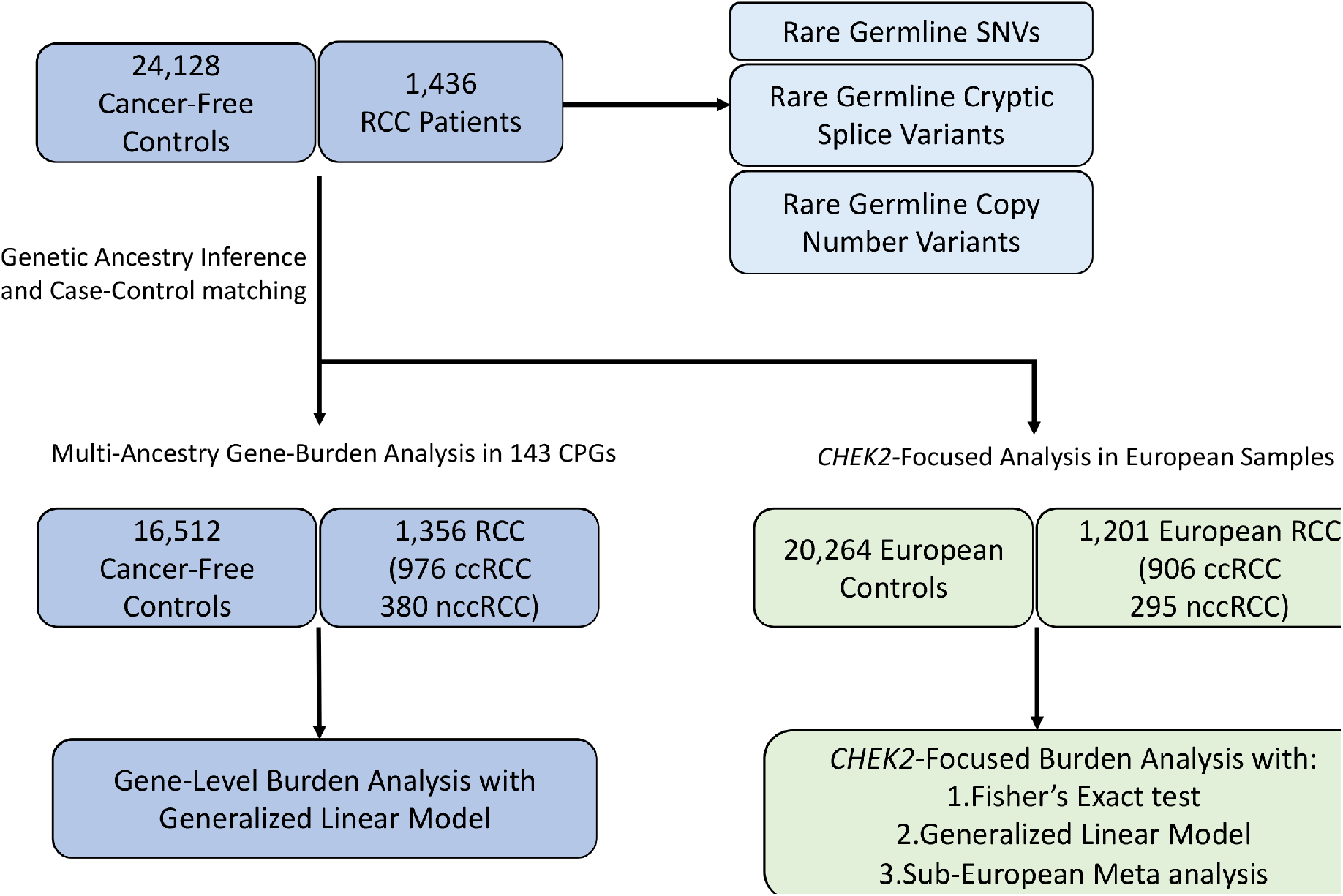
Overview of the study. Rare germline PVs, cryptic splice variants and copy number variants were characterized in 1,436 RCC patients. Gene-level burden analysis was restricted to 1,356 RCC patients ancestry-matched with 16,512 cancer-free controls after genetic ancestry inference and case-control pair-matching (Method). *CHEK2* focused analysis was restricted to 1,201 European RCC patients and 20,264 European cancer-free controls.

**Table 1.** Patient characteristics of all 1,436 renal cell carcinoma patients.

| Renal Cell Carcinoma case cohort (N=1436) |  |  |  |
| --- | --- | --- | --- |
|  |  | n | % |
| **Age at Diagnosis** | 10~19 | 1 | 0.1 |
|  | 20~29 | 10 | 0.7 |
|  | 30~39 | 46 | 3.2 |
|  | 40~49 | 191 | 13.3 |
|  | 50~59 | 397 | 27.6 |
|  | 60~69 | 469 | 32.7 |
|  | 70~79 | 249 | 17.3 |
|  | 80< | 55 | 3.8 |
|  | Age Unknown | 18 | 1.3 |
| **Self-identified Gender** | Male | 980 | 68.2 |
|  | Female | 456 | 31.8 |
| **Histology** | Clear Cell RCC | 1031 | 71.8 |
|  | Papillary RCC | 302 | 21.0 |
|  | Chromophobe RCC | 103 | 7.2 |
| **Original Study** | CHECKMATE_025 | 446 | 31.1 |
|  | TCGA_KIRC | 372 | 25.9 |
|  | TCGA_KIRP | 285 | 19.8 |
|  | ICGC_RECA_EU | 95 | 6.6 |
|  | TCGA_KICH | 76 | 5.3 |
|  | CHECKMATE_010 | 61 | 4.2 |
|  | CHECKMATE_009 | 57 | 4.0 |
|  | GENETECH_Non_ccRCC | 44 | 3.1 |
| **Inferred Ancestry** | European | 1201 | 83.6 |
|  | African | 131 | 9.1 |
|  | Admixed American | 79 | 5.5 |
|  | East Asian | 19 | 1.3 |
|  | South Asian | 6 | 0.4 |

### Cancer-Free Control Cohort

WES BAMs aligned to GRCh37 from a total of 24,128 adult unrelated individuals without known cancer diagnosis were collected from the following studies: Autism Sequencing Consortium (ASC) (Database of Genotypes and Phenotypes (dbGAP):phs000298.v4.p3), Framingham Cohort (dbGAP:phs000007.v32.p1), Multi-Ethnic Study of Atherosclerosis (MESA) Cohort (dbGAP: phs000209.v13.p3), National Heart, Lung and Blood Institute (NHLBI) GO-ESP: Lung Cohorts Exome Sequencing Project (dbGAP: phs000291.v2.p1), 1000 Genomes Project^24^. All control samples were processed using methods identical to those used for the RCC case cohort.

### Evaluation of Exome Sequencing Coverage

The average sample level sequencing coverage was calculated using Genome Analysis Toolkit (GATK)^25^ (version 3.7) tool “DepthofCoverage” to ensure that all BAMs of cases and controls had sufficient read counts to confidently call germline variants. Exome-wide mean coverage of 10X was considered the minimum acceptable coverage to ensure confident germline variant detection; samples below this threshold were excluded from our final analysis.

### Germline Variant Detection from WES Data

Germline variants were called from the BAM files using a deep learning-based variant discovery method, DeepVariant (version 0.8.0, docker: gcr.io/deepvariant-docker/deepvariant:0.8.0)^26^ that had demonstrated superior sensitivity and specificity than GATK based joint-genotyping^27,28^. All germline variants annotated with “PASS” in the FILTER column of the Variant Call Format (VCF) files were selected. Variants with low read coverage (<10 Read Depth) and low variant allele frequency (VAF <0.20) were excluded. Final sets of high-quality variants were then merged into cohort-level VCF files using GATK (version 3.7) tool “CombineVariants”. Subsequently, the ‘vt’ tool (version 3.13) was used on the cohort VCF files to normalize and decompose multiallelic variants.

### Genetic Relatedness Analysis

We performed a genetic relatedness analysis on the cohort VCF files in two steps. In the first step, we implemented GENESIS (version 2.12.0) tool PC-AiR^29^ to perform a principal components analysis (PCA) on the detected germline variants for the detection of population structure in the case and control cohort, respectively. We then used GENESIS tool “PC-Relate”^30^ implemented in “Hail”^31^ (version 0.2.11, https://github.com/hail-is/hail) to estimate kinship coefficients between every possible pair within a cohort. We removed one sample out of each pair that had a kinship coefficient above 0.125 which indicates genetic relatedness within second-degree relatives. Related samples were removed from the cohort VCF using GATK’s (version 3.7) tool “SelectVariants”.

### Genetic Ancestry Inference

First, cohort VCF files for cases and controls were combined with a cohort VCF file of 1000 Genomes Project^24^ samples (n=2,504) with known continental ancestries of Admixed American (AMR), African (AFR), European (EUR), East Asian (EAS), and South Asian (SAS), and the resulting VCF file was filtered for germline variants identified in the exome intervals well-covered (>15X read coverage) in over 95% of all samples. Next, the filtered VCF file was loaded into a matrix table using Hail (version 0.2.11, https://github.com/hail-is/hail), and rare germline variants with a cohort allele frequency below 1% and deviating from Hardy-Weinberg equilibrium (chi-squared p-value <1×10^−6^) were excluded. We next performed Linkage Disequilibrium (LD) pruning using the “ld_prune” method in Hail to prune out variants with Spearman correlation coefficient greater than 0.1 within a 1 million base pair window, and the resulting filtered germline variants were used for PCA using Hail “hwe_normalized_pca” method. Finally, Sklearn (version 0.20.0) “RandomForestClassifier” function was applied to the top 10 global principal components (PCs) of reference samples from the 1000 Genomes Project to train random forest classifiers for the 5 continental ancestries, which were used to uniformly assign continental ancestry to the cases and controls (Supplementary Figure 1A).

### Ancestry Pair-Matching of Cases and Controls

Once continental ancestry was assigned, cases and controls were divided into each continental ancestry group, and the second round of PCA was performed on each group to identify continental ancestry-specific PCs. We then used the R optmatch (version 0.9-14) package’s “pairmatch” function to identify control samples that were closest to each case based on the top 10 PCs. To ensure an equivalent representation of each ancestry group, we applied a fixed 1:12 ratio between the number of cases and controls across all continental ancestry groups, and AMR cases and controls were excluded in the gene-level burden analysis due to the limited number of AMR control samples failing to meet the case-control ratio. We also removed RCC patients with subtypes other than clear cell, papillary, or chromophobe RCC (Supplementary Figure 1B, C).

### Sub-European Ancestry Inference and Ashkenazi Jewish Inference

For sub-European ancestry inference, the same variant filtering and PCA were repeated as in the continental ancestry inference, but only using samples identified as Europeans. The top 10 PCs and sub-European ancestry labels (Finnish in Finland (FIN), Iberian Population in Spain (IBS), Toscani in Italia (TSI), Utah Residents with Northern and Western European Ancestry (CEU), and British in England and Scotland (GBR)) from the 1000 Genomes European samples were used to train a random forest classifier. The classifier was applied to the case and control samples to infer sub-European ancestry.

Since Ashkenazi Jewish (ASJ) individuals were unable to be identified using the above approach, we used SNPweights^32^ software with pre-calculated SNP weights from Ashkenazi Jewish reference samples to identify ASJ individuals (Supplementary Figure 3). Samples with ASJ proportion >0.5 were defined as ASJ. In the end, European cases and controls were divided into Northwestern Europeans (including CEU and GBR), Southern Europeans (IBS and TSI excluding ASJ), Finnish, and Ashkenazi Jewish.

### Functional and Clinical Annotation and Prioritization of Germline Variants

Germline variants in the cohort VCF files were annotated using Variant Effect Predictor (VEP, version 104.3)^33^ with dbNSFP^34^ (version 4.1a, GRCh37) and ClinVar^35^ (Release 20220829, GRCh37) plug-ins. A curated list of 143 CPGs (eTable 2) was used to identify candidate rare (Minor Allele Frequency (MAF) <1%) germline PVs. All identified variants were then classified into five categories: benign, likely benign, variants of unknown significance, likely pathogenic, and pathogenic, using the American College of Medical Genetics (ACMG) classification^36^ provided by VarSome^37^ website (accessed between September-November 2022). Variants classified as likely pathogenic or pathogenic are collectively referred to as pathogenic variants (PVs). The presence of the identified PVs was further validated by manual examination of the BAM files using Integrative Genomics Viewer (IGV, version 2.11.1).

### Gene-level Burden Analysis with a Generalized Linear Model

To perform gene-level burden analysis, a null model based on a generalized linear model (GLM), as implemented in the Python “statsmodel” library (version 0.13.2)^38^ was first constructed using the top 10 global PCs from ancestry inference as covariates and RCC case status as the dependent variable. For each gene with at least one pathogenic variant in RCC cases or controls, a corresponding extended model incorporating a burden indicator variable representing the presence of a pathogenic variant in the gene for every sample was constructed. A likelihood ratio test was then performed between the null model and each extended model, and the resulting test statistics were adjusted for the false discovery rate (FDR) using the Benjamini-Hochberg procedure with FDR=0.05. The burden of three low-penetrance *CHEK2* variants defined by a recent study^12^ - *CHEK2* p.Ile157Thr, p.Ser428Phe, and p.Thr476Met – were evaluated separately from the pathogenic LOF variants identified in *CHEK2*.

### Statistical Analysis and Data Visualization

Odds ratios, 95% confidence intervals, and p-values for two-sided Fisher’s Exact test were computed as implemented in the exact2×2 R package. Adjusted p-values (q-values) were computed based on the Benjamini-Hochberg procedure with FDR=0.05. A one-way ANOVA test was run using the “f_oneway” function from Python “scipy” library (version 1.5.2)^39^, and *post hoc* pairwise comparisons were performed where applicable using the “pairwise_tukeyhsd” function from Python “statsmodels” library. Sample proportion confidence intervals were calculated using the R “prop.test” function. For the meta-analysis, association statistics from sub-European groups were combined using a fixed-effects meta-analysis implemented using the “metafor” R package^40^. All figures were generated using Python “Seaborn” (version 0.11.0, Waskom M) and “Matplotlib” (version 3.3.2) packages and were further refined using Adobe Photoshop 2021.

The commutation plot (Figure 4) summarizing the germline and somatic variants in RCC cases was generated using Python “CoMut” package^41^ (version 0.0.3, https://github.com/vanallenlab/comut).

### Identification of Somatic Variants and Copy Number Events

For carriers of germline pathogenic variants in *VHL, MET, FH*, and *CHEK2*, somatic variants data provided from TCGA and ICGC were downloaded from the GDC data portal (https://portal.gdc.cancer.gov/) and ICGC data portal (https://dcc.icgc.org/) respectively (accessed: Oct. 2022) and was analyzed to identify somatic variants and copy number alterations involving the genes of interest. For somatic variants, LOF truncating variants including frameshift, splice site, or nonsense variants as well as missense variants with oncogenic annotation from OncoKB were included. For samples from CheckMate studies, somatic variants and copy number alterations were identified with the CGA WES Characterization pipeline (https://github.com/broadinstitute/CGA_Production_AnalysisPipleline), which detects, filters, and annotates somatic variants and copy number alterations. The CGA pipeline employs the following tools: MuTect^42^, ContEst^43^, Strelka^44^, Orientation Bias Filter^45^, DeTiN^46^, AllelicCapSeq^47^, MAFPoNFilter^48^, RealignmentFilter, ABSOLUTE^49^, GATK^25^, PicardTools (https://software.broadinstitute.org/gatk/documentation/tooldocs/4.0.1.0/picard_fingerprint_CrosscheckFingerprints.php), Variant Effect Predictor^33^,and Oncotator^50^. Samples were excluded for quality control as previously described^51^. Briefly, tumor samples with ≥ 5% contamination^43^, normal samples contaminated with ≥ 20% tumor nuclei^46^, samples with poor sequencing coverage (tumor mean target coverage < 25x, normal mean target coverage < 15x), and samples identified as a tumor/normal swap based on the copy number profile (lane mix-up > 1) were excluded from further analyses. To assess tumor purity, both ABSOLUTE^49^ and FACETS^52^ were used. ASBOLUTE solutions were reviewed for tumor samples with < 20% estimated purity from FACETS and samples were subsequently excluded if < 20% purity was estimated from ABSOLUTE. To ensure adequate power for detecting somatic variants, a power calculation for each tumor sample was performed as previously described and samples with < 80% power were excluded^53^.

### Identification and Validation of Cryptic Splice Variants

SpliceAI (version 1.3.1, https://github.com/Illumina/SpliceAI)^54^ was used to identify cryptic splice variants among the called germline variants detected by DeepVariant. The “SpliceAI” INFO column provided Delta scores ranging from 0 to 1 for the probability of acceptor gain/loss and donor gain/loss. The maximum Delta score was defined as the “SpliceAI” score in the annotated cohort VCF files, and the putative cryptic splice variants were further categorized according to the official cutoff recommendations: 0.2 (high recall), 0.5 (recommended), and 0.8 (high precision). Rare germline variants with SpliceAI score over 0.5 were defined as putative cryptic splice variants. For samples carrying a cryptic splice variant identified from SpliceAI, available tumor or germline mRNA BAM files were manually reviewed using IGV to visualize and evaluate their splicing patterns.

### CNV Detection from Whole Exome Sequencing Data

We applied GATK-gCNV to detect rare germline CNVs from exome sequencing data^55^. To minimize the discrepancy among the different exome sequencing baits used for different sources, GATK (version 4.1.9.0) tool “CollectReadCounts” was used to gather read counts on the 8441 sequencing bait regions unique to 7 major capture kits, and PCA was run to make batches of samples sequenced using the same sequencing bait (Supplementary Figure 5A). From each identified batch, germline CNVs were detected using GATK-gCNV^55^ following the best practices on the Terra platform (https://app.terra.bio/#workspaces/help-gatk/Germline-CNVs-GATK4). The detected CNVs were harmonized and filtered using the gCNV filtering R scripts downloaded from the gCNV repository (https://github.com/theisaacwong/talkowski/tree/master/gCNV), which combines CNVs from multiple batches and identifies high-quality rare CNVs determined by assessing 3 quality control measures – Quality Score (QS), the rarity of a CNV (PASS_Freq, for site frequency of 1% or less), and the number of samples carrying a rare CNV (PSS_Sample, Samples carrying more than 100 unfiltered or more than 10 filtered calls are excluded).

## RESULTS

### Patient Characteristics of RCC Discovery Case Cohort

We collected whole-exome sequencing (WES) data from 1,436 RCC patients unselected for earlier age of disease onset or positive family history from 8 independent RCC studies (Figure 1, Table 1). 71.8% of patients had clear cell RCC (ccRCC, n=1,031), while the rest had non-clear cell RCCs (nccRCC), including papillary (n=302, 21.0%) and chromophobe RCC (n=103, 7.2%). Broad continental-level genetic ancestry inference (Supplementary Figure 1) identified most of the cohort as being of predominantly European ancestry (83.6%, n=1,200) followed by African (9.1%, n=131), Admixed American (5.6%, n=80), East Asian (1.3%, n=19), and South Asian (0.4%, n=6).

### Prevalence of Rare Germline Pathogenic Variants in ccRCC and nccRCC

We first evaluated rare (MAF<1%) germline variants that met existing clinical interpretation guidelines^36^ as pathogenic or likely pathogenic in 1,031 ccRCC patients (Figure 2). In known RCC risk genes, we identified rare germline PVs in *VHL* (n=4, 0.38%, 95% CI: 0.12-1.1%), *BAP1, MITF* (n=3 each, 0.29%, 95% CI: 0.075-0.92%), *FLCN, FH*, and *SDHD* (n=1 each, 0.097%, 95% CI: 0.0051-0.63%). When evaluating DNA damage repair (DDR) genes (eTable 3), we identified 52 ccRCC patients that harbored rare germline PVs in homologous recombination or Fanconi Anemia genes such as *CHEK2, RECQL4, FANCA*, or *BRCA1/2* (5.04%, 95% CI: 3.82-6.60%), 26 in base excision repair genes *MUTYH* and *NTHL1* (2.52%, 95% CI:1.69-3.73%), 8 in nucleotide excision repair genes; *ERCC1, ERCC2, ERCC3, XPA*, and *XPC* (0.77%, 95% CI:0.36-1.59%), and 2 in mismatch repair genes *MLH1* and *PMS2* (0.19%, 95% CI:0.033-0.79%). Overall, 131 ccRCC patients carried one or more heterozygous rare germline PVs (12.71%, 95% CI:10.77-14.93%; eTable 4); 13 in previously established RCC risk genes (1.26%, 95% CI: 0.71-2.21%), 86 in DDR genes (8.34%, 95% CI:6.76-10.24%), and 37 in rest of the germline cancer predisposition genes (CPGs, 3.59%, 95% CI:2.57-4.96%). Among these, 9 ccRCC patients carried rare germline PVs in two different CPGs (0.87%, 95% CI: 0.43-1.71%; eTable 6).

**Figure 2.**
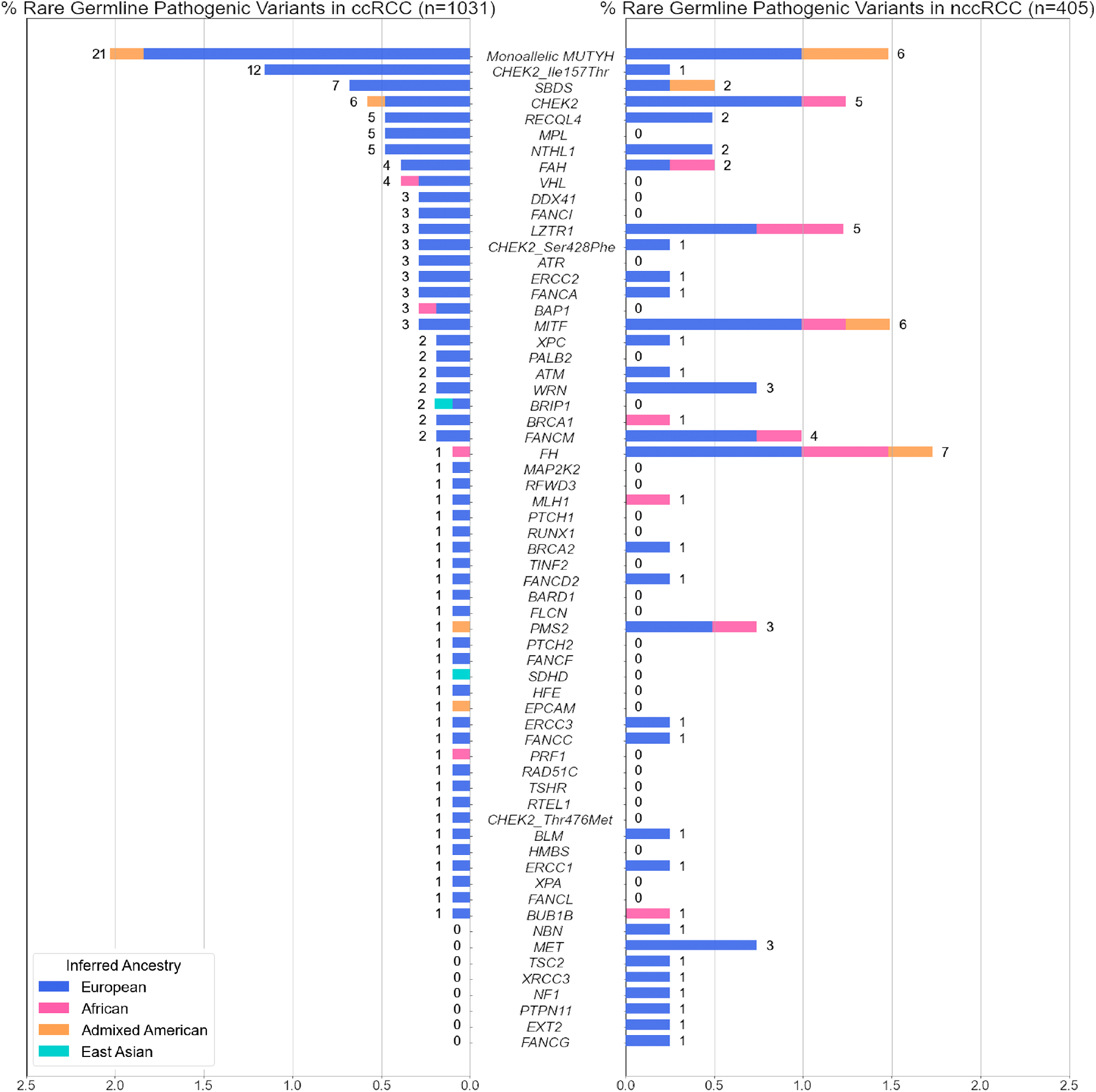
Rare germline pathogenic variants identified in the 1,436 RCC patients. The proportion of ccRCC or nccRCC patients carrying rare (MAF <1%) germline pathogenic variants in one of the 143 germline cancer predisposition genes tested. 3 known *CHEK2* low-penetrance variants were treated separately from the rest of the PVs in *CHEK2*. Numbers next to the bars indicate counts of RCC patients carrying PVs in each gene.

In parallel, we also characterized rare germline PVs in 405 nccRCC patients (Figure 2). Rare germline PVs were found in the following known kidney cancer risk genes: 7 in *FH* (1.72%, 95% CI:0.76-3.69%), 6 in *MITF* (1.48%, 95% CI:0.60-3.36%), 3 in *MET* (0.74%, 95% CI:0.19-2.33%), and 1 in *TSC2* (0.25%, 0.013-1.59%). Regarding DDR genes, 24 germline PVs were detected in homologous recombination or Fanconi Anemia genes (5.93%, 95% CI:3.91-8.81%), 8 in base excision repair genes with PVs in ccRCC - *MUTYH* and *NTHL1* (1.98%, 95% CI:0.92-4.01%), and 4 each in mismatch repair and nucleotide excision repair genes (0.99%, 95% CI:0.32-2.69% each). Altogether, one or more rare pathogenic germline PVs were detected in 68 nccRCC patients (16.79%, 95% CI:13.35-20.87%; eTable 5) – 17 in known RCC risk genes (4.20%, 95% CI: 2.54-6.77%), 39 in DNA damage repair genes (9.63%, 95% CI:7.02-13.03%), and 13 in other CPGs (3.21%, 95% CI:1.79-5.57%). Four patients were identified with rare germline PVs in two different CPGs (0.99%, 95% CI:0.32-2.69% eTable 6). Thus, the relatively higher proportion of patients with identified germline PVs in DDR genes, which was mainly driven by rare germline PVs in *CHEK2* and *MUTYH* (3.90%, n=56/1436 across RCC subtypes, 95% CI: 2.98-5.07%) was consistent with observations in the pan-RCC patients from previous RCC studies^6,8-10^.

### Gene-Level Enrichment of Rare Germline Pathogenic Variants in ccRCC and nccRCC Patients

To investigate whether the identified PVs predispose individuals to an increased risk of RCC, we performed genetic ancestry inference and case-control pair matching to link 1,356 RCC patients (of the original 1,436; 94.4% of the RCC cohort) with 16,512 cancer-free controls (Method; Supplementary Figure 1) and compared the gene-level burden of rare germline PVs in ccRCC (n=976) and nccRCC (n=380) separately against the cancer-free controls. To further account for residual population stratification not captured in our ancestry matching procedure, we conducted gene-level burden analysis for each gene using a generalized linear model (GLM) that accounts for continental ancestry (Methods). As expected, ccRCC patients exhibited a significantly higher enrichment of germline PVs in *VHL* compared to the ancestry-matched controls (OR: 39.1, 95% CI: 7.01-218.07, p-value:4.95e-05, q-value:0.00584), and in the low-penetrance *CHEK2* p.Ser428Phe variant (OR: 31.96, 95% CI: 6.23-163.89, p-value:0.000385, q-value:0.0227) after multiple-hypothesis correction (Table 2). ccRCC patients also carried a nominally higher frequency of PVs in the two other known kidney cancer risk genes, *BAP1* and *SDHD*, as well as in the common low penetrance *CHEK2* p.Ile157Thr variant (p-values <0.05, q-values > 0.05; eTable 7).

**Table 2.** Gene-level burden analysis of clear cell and non-clear cell RCCs. Test statistics from the generalized linear model for genes with nominal enrichment. Genes with adjusted p-value less than 0.05 were considered significant and highlighted. A table of results for all genes tested can be found in eTables 7 and 8

|  | Gene/Variant | FDR_Adjusted_p-value | p-value | Odds_ratio | OR_95CI_low | OR_95CI_high | case_PV_carrier | control_PV_carrier |
| --- | --- | --- | --- | --- | --- | --- | --- | --- |
| Clear Cell RCC | *VHL* | 0.00584 | 4.95E-05 | 39.1 | 7.01 | 218.07 | 4 | 2 |
|  | *CHEK2_Ser428Phe* | 0.0227 | 0.000385 | 31.96 | 6.23 | 163.89 | 3 | 3 |
|  | *SDHD* | 0.506 | 0.0141 | 1.72E+10 | 0 | inf | 1 | 0 |
|  | *BAP1* | 0.526 | 0.0314 | 5.57 | 1.44 | 21.52 | 3 | 9 |
|  | *CHEK2_Ile157Thr* | 0.526 | 0.0356 | 2.04 | 1.11 | 3.73 | 12 | 122 |
| Non Clear Cell RCC | *FH* | 1.83E-06 | 1.55E-08 | 77.9 | 18.68 | 324.97 | 6 | 3 |
|  | *MET* | 2.07E-05 | 3.50E-07 | 1.98E+11 | 0 | inf | 3 | 0 |
|  | *LZTR1* | 0.244 | 0.00619 | 4.96 | 1.92 | 12.85 | 5 | 42 |
|  | *PMS2* | 0.653 | 0.0241 | 5.58 | 1.65 | 18.84 | 3 | 25 |
|  | *MITF* | 0.653 | 0.0277 | 3.38 | 1.34 | 8.53 | 5 | 62 |
|  | *CHEK2_Ser428Phe* | 0.707 | 0.036 | 27.73 | 2.78 | 276.8 | 1 | 3 |
|  | *EXT2* | 0.79 | 0.0483 | 22.36 | 2 | 250.27 | 1 | 2 |

For nccRCC, patients carried a significantly higher prevalence of germline PVs compared to the controls in *FH* (OR: 77.9, 95% CI: 18.68-324.97, p-value:1.55e-08, q-value: 1.83e-06) and *MET* (OR: 1.98e11, 95% CI: 0-inf, p-value: 2.07e-05, q-value: 3.50e-07). *LZTR1, PMS2, MITF, EXT2*, and *CHEK2* p.Ser428Phe also exhibited nominal enrichment of germline PVs but did not pass multiple hypothesis correction (p-values <0.05, q-values > 0.05, Table 2). In contrast to prior studies^9,11,12^, LOF variants in *CHEK2* were not significantly enriched in ccRCC after excluding low-penetrance variants (OR: 1.01, CI: 0.41-2.52; p-value of 0.980, q-value: 0.997) or nccRCC (OR: 2.82, CI: 1.13-7.05; p-value of 0.0536, q-value: 0.79). No other DDR genes were enriched with PVs in ccRCC or nccRCC patients compared to the cancer-free controls (eTables 7 and 8).

### Evaluation of *CHEK2* in RCC Risk Via Accounting for Fine-Scale Genetic Differences in European Sub-populations

The three main germline PVs identified in *CHEK2* in our study – c.1100del, Ser428Phe, and Ile157Thr – are all founder variants from different European sub-populations, with substantial variation in population minor allele frequencies across different European populations (eTable 9)^56^. These variants were also identified at different frequencies in our cases and controls of different sub-European ancestries (Supplementary Figure 2). Therefore, to explore whether subtle ancestry differences in European populations may have confounded *CHEK2* rare variant analyses, we performed three additional *CHEK2* burden analyses restricted to European samples to evaluate the impact of addressing fine-level population stratification on the role of *CHEK2* as an RCC risk gene: (1) a Fisher’s Exact-based association study on all Europeans pooled together; (2) a GLM-based burden-analysis using the top 10 genetic principal components from a European-only principal components analysis; (3) a meta-analysis combining test statistics from different sub-European populations (Figure 3; Methods). *CHEK2* germline LOF PVs did not demonstrate enrichment in ccRCC cases in all three tests, though they exhibited a nominal enrichment in nccRCC only in the meta-analysis (OR: 3.51, 95% CI:1.10-11.10, combined p-value: 0.0330, Figure 3A). In contrast, only ccRCC patients exhibited a nominally higher burden of *CHEK2* p.Ile157Thr variant in the meta-analysis (OR: 1.84, 95% CI:1.00-3.36, combined p-value: 0.0486) (Figure 3B). Finally, this expanded statistical framework demonstrated that the *CHEK2* p.Ser428Phe variant that was significantly enriched in the above multi-ancestry GLM-based burden analysis were only modestly enriched in these ccRCC patients (OR:5.20, 95% CI:1.00-26.40, combined p-value:0.0449), reflecting the localized burden of the variant in the inferred Ashkenazi Jewish RCC individuals in cases and cancer-free controls (Figure 3C). Taken together, the results demonstrate that the risk assessment of *CHEK2* germline variants requires careful consideration of population stratification due to the varying frequency of founder variants in this gene.

**Figure 3.**
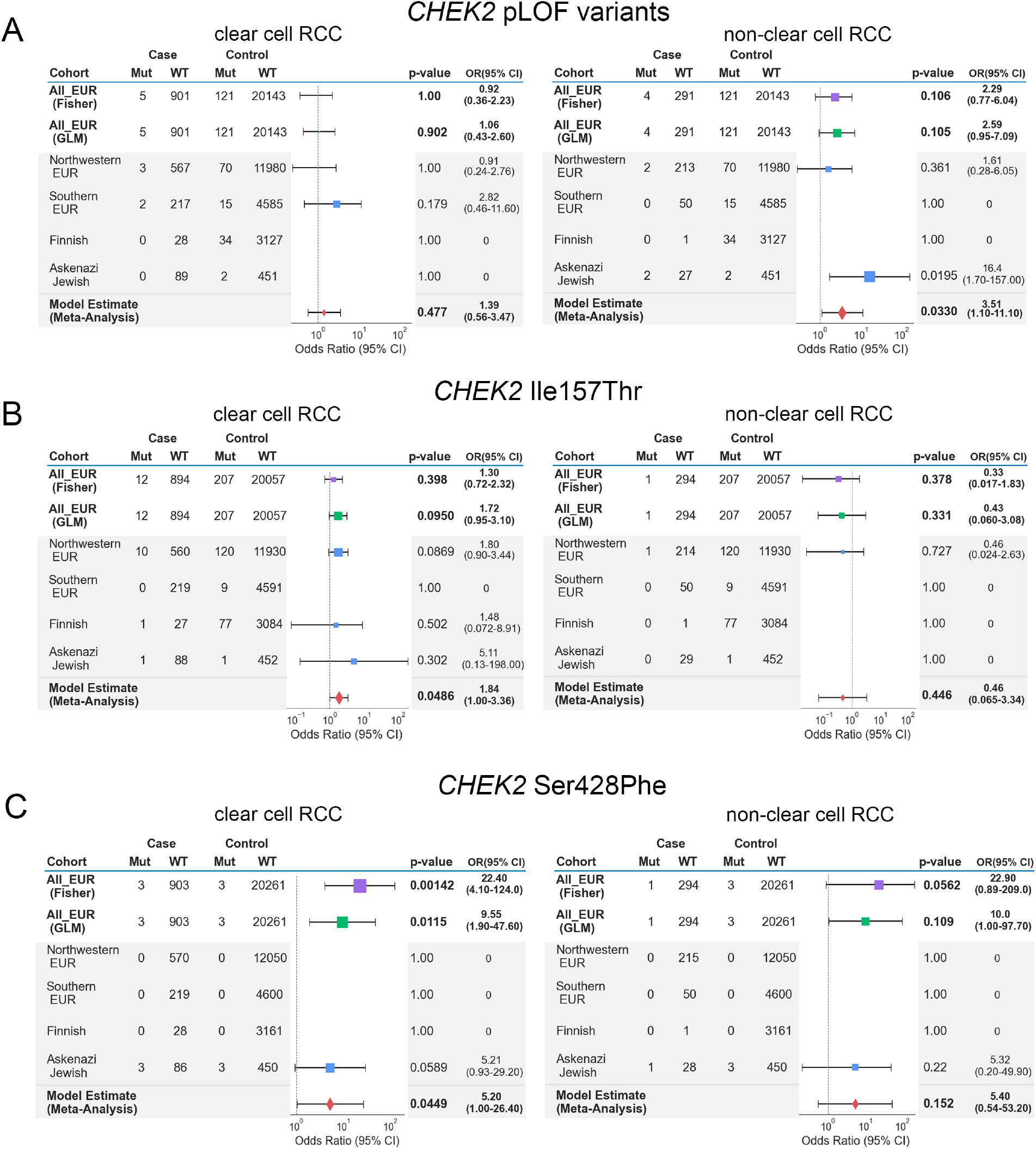
Meta-Analysis of *CHEK2* risk in ccRCC and nccRCC in European samples. Tables and forest plots summarize the model estimates summary statistics from the sub-European meta-analysis for *CHEK2* germline variants. The area of squares is proportional to the -log10 of the p-values, and the horizontal bars indicate 95% confidence intervals for the estimated odds ratio. Test statistics from Fisher’s Exact and GLM tests were plotted for comparison. A) Summary tables for *CHEK2* LOF excluding the low-penetrance variants B)Summary tables for *CHEK2* founder variant p.Ile157Thr c)Summary tables for *CHEK2* founder variant p.Ser428Phe

### Prevalence of Somatic Second-Hit Variants in Carriers of Germline Pathogenic Variants

To further clarify the potential roles of the rare germline PVs identified in these analyses, we next investigated the available tumor samples from the RCC patients in our study to identify somatic events (truncating somatic variants or copy number alterations; Methods) accompanying the germline PVs identified in our analyses (Figure 4A; eTable 10). Tumors from ccRCC patients carrying germline PVs in *VHL* had a somatic copy number deletion in chromosome 3 spanning *VHL* (n=3/4, 75.0%, 95% CI: 21.94-98.68%).^57^ Regarding nccRCC, all 3 carriers of germline *MET* PVs were patients with type 1 papillary RCC (pRCC) whose tumors had somatic copy number gains at chromosome 7 (spanning the *MET* locus), while 87.5% (n=7/8, 95% CI: 46.68-99.34%) of patients with germline PVs in *FH* were from type 2 pRCC whose tumors often had somatic variants or copy number deletions in *FH* (n=4/7, 57.1%, 95% CI: 20.24-88.19%). Overall, 10 patients (66.7%, 95% CI: 38.69-87.01%) carrying germline PVs in *FH, MET*, or *VHL* harbored identifiable secondary somatic events in the same genes, further indicating the importance of these genes in the RCC oncogenesis. In contrast, patients carrying *CHEK2* germline variants were not limited to a specific RCC subtype, and only 7 of 23 (30.4%, 95% CI: 14.06-53.01%) RCC patients with germline variants in *CHEK2* had secondary somatic variants in *CHEK2* (1 patient with both somatic variant and copy number deletion in *CHEK2*; 6 patients with *CHEK2* copy number deletions).

**Figure 4.**
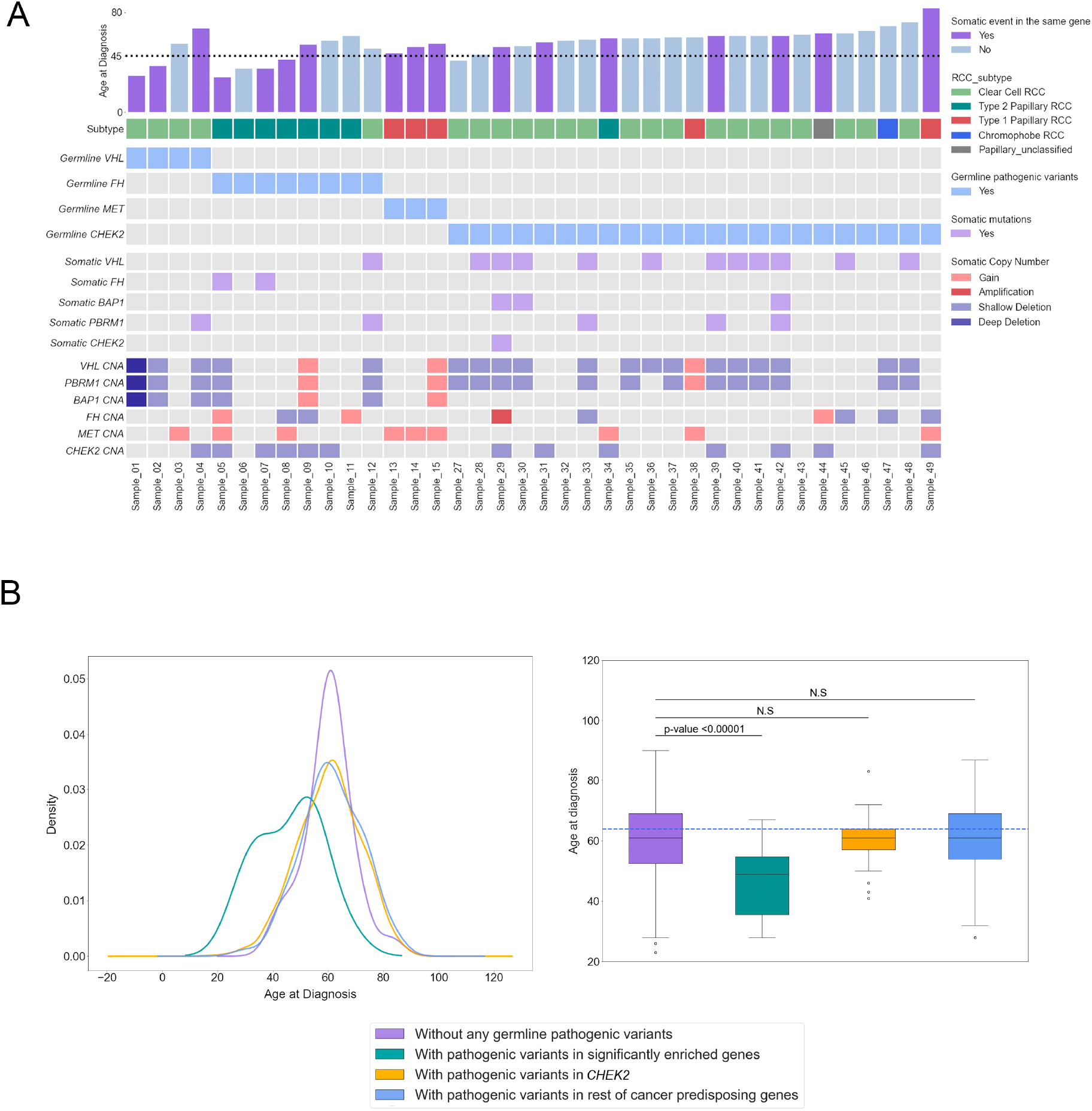
Somatic events in the carriers of pathogenic variants in genes with enrichment. A)Comutation plot summarizing germline and somatic variants in RCC patients with germline pathogenic variants in the three significantly enriched genes as well as *CHEK2*. For the somatic events, only variants in relevant genes are featured. The dotted horizontal line on the bar plot indicates the age of onset at 45. CNA indicates copy number alterations B)Density plot (left) and Box plot (right) for the distribution of age of disease onset for the different pathogenic variant carrier groups. Adjusted p-value was calculated after a one-way ANOVA with *post hoc* Tukey’s HDS test.

### Age of RCC Presentation for Carriers of Germline Pathogenic Variants in RCC Risk Genes

Rare germline PVs in genes known to cause hereditary cancer syndromes have been associated with an earlier onset of disease in different cancers including RCC^58-62^, and detection of these variants with genetic testing can guide clinical management in at elevated genetic risk for cancer^63^. To further characterize the clinical impact of the rare germline PVs identified in the genes with a significantly higher burden of PVs in RCC patients, we compared the age of disease onset between the groups defined by genetic status: (1) patients carrying germline PVs in the known RCC risk genes; *FH, MET*, and *VHL*; (2) patients carrying germling PVs in *CHEK2*; (3) patients carrying germline PVs in other CPGs without enrichment; (4) patients carrying no germline PVs (Figure 4B; eTable 11). The carriers of rare germline PVs in *FH, MET*, and *VHL* presented with disease at a significantly earlier age compared to the other three groups (Mean: 46.0, Median: 49.0 years old, Tukey *post hoc* adjusted p-values < 0.01 for all three pairwise comparisons), and patients with germline and somatic biallelic events presented with disease at an earlier age (n=10/15, Mean: 44.6, Median: 44.5 years old, Figure 4A). However, the age of disease onset for the patients carrying germline *CHEK2* PVs showed no evidence of age difference from that of patients carrying no germline PVs (Mean: 60.1, Median: 61 vs Mean: 60.2, Median: 61 years old, Tukey *post hoc* adjusted p-value =1.0) or patients carrying PVs in the rest of CPGs that did not exhibit enrichment in RCC (Mean: 60.1, Median: 61 vs Mean: 61.3, Median: 61 years old, Tukey *post hoc* adjusted p-value =0.949). Similarly, RCC patients carrying both germline and somatic variants in *CHEK2* did not present at an earlier age of onset compared to patients without any germline PVs (n=7/23, Mean: 62.1, Median: 61.0 years old). These results, taken together with the only modest enrichment of germline PVs in *CHEK2*, suggests caution for considering *CHEK2* as a RCC predisposition gene.

### Identifying Additional Forms of Inherited Genomic Alterations in RCC Risk Genes

While 13.9% (n=199/1436, 95% CI: 12.13-15.78%) of total RCC (ccRCC and nccRCC) patients carried rare germline PVs in CPGs, only 15 RCC patients (1.04%, 95% CI: 0.61-1.76%) harbored germline PVs in known RCC risk genes (*FH, MET*, and *VHL*), but the rest of the CPGs did not exhibit increased burden of PVs in RCC patients in our analyses and thus are of uncertain biological significance in RCC pathogenesis. Thus, we hypothesized that RCC risk genes may also be disrupted through mechanisms that can escape the detection of conventional germline variant detection methods commonly used in clinical and research contexts, such as cryptic splice variants outside of the canonical splice sites and germline copy number variants (CNVs). Using existing computational methods to predict cryptic splice variants, we identified 109 candidate rare germline cryptic splice variants in CPGs in 102 RCC patients (Supplementary Figure 4A; eTable 12; Methods). Of these, 86 patients had tumor and/or germline mRNA sequencing data available to validate these predicted splice variants.

The available RNA sequencing data showed no evidence of aberrant splicing for 82 variants (95.35%, 95% CI: 87.87-98.50%). However, two cryptic splice variants in two RCC risk genes, *TSC1* and *SDHA*, demonstrated a clear pattern of aberrant splicing (Figure 5). The cryptic splice variant in *TSC1* in a chromophobe RCC patient changed antisense cytosine upstream of a splice donor motif to thymine, leading to complete exon-skipping of exon 21. In the papillary RCC patient with a variant in *SDHA*, the cryptic splice variant introduced a cryptic donor motif inside exon 13 and removed 15 amino acids at the end of the exon. Two other cryptic splice variants in *TP53* and *LZTR1* showed aberrant splicing, but the number of splice junction reads was too low to confidently conclude them as clear splice variants (Supplementary Figure 4B; Methods).

**Figure 5.**
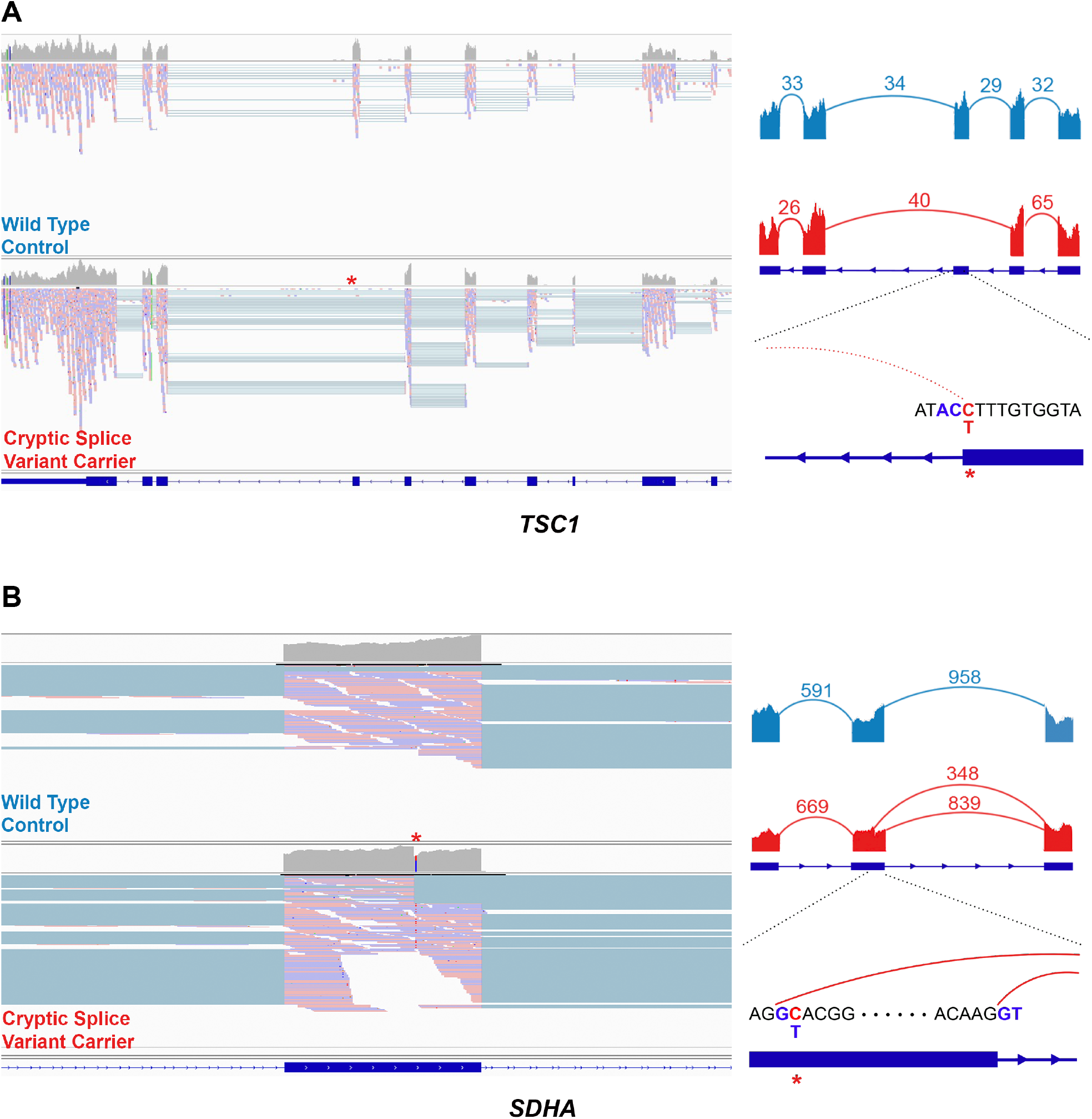
Example cryptic splice variants in established RCC risk genes. A) Left: IGV screenshot of the tumor mRNA sequencing data of the wildtype control (above) and the carrier of cryptic splice variant (bottom). Right: Sashimi plot showing the pattern of splicing with the numbered split junction reads. A)Disruption of splice donor motif led to complete exon skipping in *TSC1* B)The cryptic splice variant in *SDHA* introduced a new splice donor motif GT inside an exon

We next evaluated germline CNVs (Methods). Collectively, we identified 2,503 high-quality rare germline CNVs in 888 RCC samples (1,211 deletions and 1,292 duplications; Supplementary Figure 5B-C; Methods). Of these, 18 heterozygous CNVs in 18 (1.25%, 95% CI: 0.77-2.02%) RCC patients affected 14 CPGs including RCC risk genes *FH, VHL*, and *SDHA* (Figure 6A; eTable 13). For example, we found a ccRCC patient harboring a deletion spanning part of the last exon of *VHL* (Chr3:10191124-10192282, GRCh37, Figure 6C), and another deletion was identified in a papillary RCC patient overlapped the last 761bp of *FH* (Chr1:240070386-241661618, Figure 6D). We also identified a large 215 kbp deletion completely spanning *SDHA* in a different papillary RCC patient (Chr5:139251-354374, Figure 6B), which disrupted a region similar to a variant previously described in the gnomAD structural variant (SV) database (gnomAD ID: DEL_5_54065, Chr5:135575-308762)^64^. Thus, by characterizing underappreciated variant types such as cryptic splice and CNVs, we identified 6 additional putative pathogenic variants in the established RCC risk genes, increasing the diagnostic yield of rare germline PVs in risk genes from 2.1% (n=30/1436, 95% CI: 1.44-3.01%) to 2.5% (n=36/1436, 95% CI: 1.79-3.49%).

**Figure 6.**
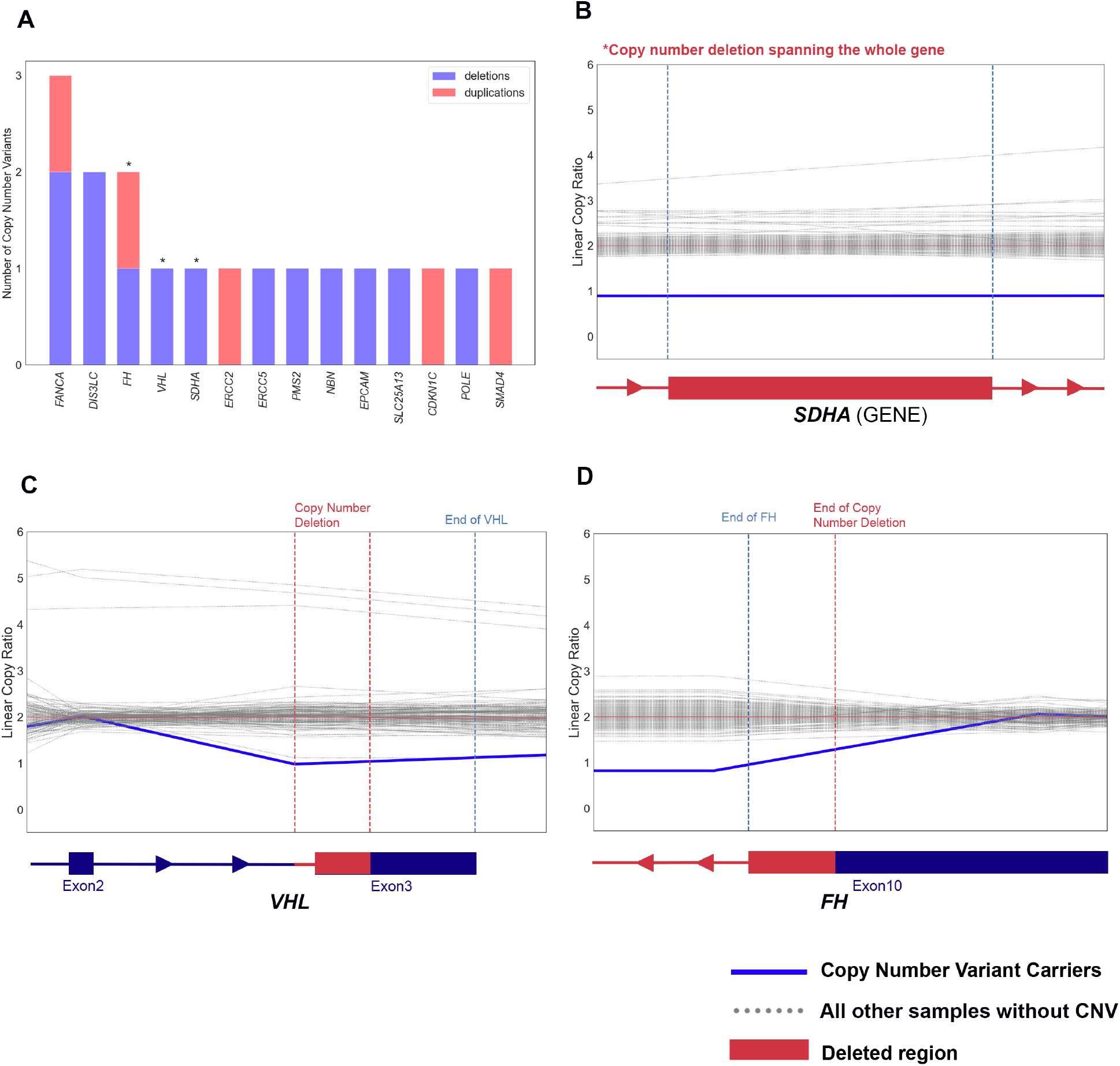
Example rare germline copy number variants (CNVs) in established cancer-predisposing genes. A) Bar-plot summarizing the counts of deletions and duplications including the germline cancer predisposing genes B-D) Denoised linear copy ratio (DCR) plot indicating heterozygous copy number deletion with copy ratio of 1 for the carriers of CNV. The blue line indicates DCR for the CNV carrier, and the dotted grey lines indicate DCR for the rest of wild type samples.

## DISCUSSION

Thus far, fourteen genes in the HIF, PI3K/mTOR, chromatin regulation, and cell cycle pathways have been identified as established RCC risk genes^65^, but the estimated high genetic heritability of RCC is not fully explained by germline PVs detected in these genes. Multiple recent studies have built on these foundational discoveries to report frequencies of rare germline PVs in DDR genes in large RCC cohorts. However, analyses with proper cancer-free controls and statistical models accounting for population structure are necessary to determine whether these PVs are significantly enriched in RCC populations. Attempts to demonstrate association by comparing frequencies of PVs in RCC cases against public databases such as ExAC^66^ or gnomAD^56^ are fraught with major technical limitations, including that the samples are likely not sequenced using the same sequencing platform, variants were not called using the same variant discovery pipeline and were not processed identically, and cases and controls were not ancestry-matched to ensure a robust statistical comparison. Recently, two studies leveraged case-control approaches to report an association of rare germline PVs in *CHEK2* with an elevated risk of RCC^11,12^. However, both studies treated different subtypes of RCC together as a single phenotype and included *CHEK2* variants with distinct population properties. To address the gaps of knowledge in the field, we performed histology-specific and case-control analyses of rare germline PVs in RCC, finding that PVs in the three significantly enriched genes – *VHL, MET*, and *FH* – had no significant overlap in different RCC subtypes. This molecular difference was also clearly demonstrated when we investigated companion somatic variants and copy number events stratified by histological subtype.

Furthermore, while adjusting for finer-scale population stratification is the current standard in GWAS studies, many case-control rare germline variant studies in cancer (including most RCC studies) only adjusted for population stratification by limiting their association analysis to cases and controls of European descent under the assumption that individuals of broad continental European ancestry would have a similar genetic background. Consistent with prior observations that population stratification can confound rare variant association studies in other disease contexts^67-70^, we found that a standard Fisher’s Exact test-based association model without addressing finer-scale population stratification can lead to a false association in this context, especially for *CHEK2* that has several founder variants with varying population frequency within Europe. For example, the most well-studied *CHEK2* c.1100del variant’s population frequency substantially varies in different regions of Europe, ranging from ∼0% in Spain to 1.6% in the Netherlands and has a relatively lower population frequency in North America compared to Europe^71-77^. These factors may explain why in our study, the *CHEK2* variant was detected in only 0.35% of RCC cases, where most patients were from the U.S. and patients were unselected for family history. Meanwhile, this variant was identified in more than 1% of RCC cases in a UK-based RCC association study^11^ and significant enrichment of *CHEK2* germline PVs was reported in studies with patients selected for positive family history^9^ or positive *CHEK2* germline variant carrier status in panel sequencing^12^. The variation in population frequency of *CHEK2* variants combined with prior studies lacking robust genetic ancestry inference and case-control matching may partially explain the disagreeing risk assessment within and across cancer types for this gene^78^. Our result warrants caution against the commonly used practice of treating all “white” or “Caucasian” individuals with predominantly European ancestry as one group in cancer genetic studies. This practice can substantially confound association studies, particularly for studies including participants from the U.S. population which consists of different European ancestry groups as well as non-European ancestry groups.

Furthermore, in this study, RCC patients with germline PVs in *CHEK2* did not have frequent secondary somatic events, whereas tumors from carriers of PVs in *bona fide* RCC predisposition genes like *FH, MET*, or *VHL* largely exhibited secondary somatic events in these genes. In addition, in this cohort of RCC patients unselected for the age of disease onset, RCC patients with germline PVs in *CHEK2* did not exhibit earlier age of RCC onset, contrasting the relatively early age of breast cancer onset for germline *CHEK2* variant carriers reported in several studies^71,73,79-81^, which suggests uncertainty regarding the role of *CHEK2* germline PVs in RCC risk. Unlike in breast cancer, the biological role of DDR genes in Fanconi anemia or homologous recombination pathways such as *CHEK2* has not definitively been demonstrated in RCC, and we did not identify an enrichment of germline PVs in other DDR genes besides *CHEK2*. Given these observations, we suggest caution in including *CHEK2* or any other DDR genes as RCC risk genes. Critically, our analysis does not preclude the association of *CHEK2* with RCC but advises for addressing population stratification in larger cohorts that include different sub-European ancestry groups is warranted to clarify the role of this gene in RCC risk and heritability.

Moreover, the general focus on germline small nucleotide variants and small indels in coding sequences in prior studies may limit our understanding of potential PVs in established RCC risk genes or other candidate genes. Thus, we also investigated rare germline cryptic splice variants and germline CNVs that have not been well-characterized in RCC or cancer germline studies. Cryptic splice variants can introduce or remove splicing donor or acceptor motifs inducing aberrant mRNA splicing and loss of protein function^82,83^. However, they can be easily overlooked as non-pathogenic because they are usually annotated as non-truncating when using conventional annotation approaches. In this study, we identified two rare germline cryptic splice variants which induced aberrant splicing in RCC risk genes *SDHA* and *TSC1*, which appear to reduce wild-type transcript abundance based on our investigation of matched transcriptome sequencing data. To our knowledge, this is the first description of germline cryptic splice variants in RCC and may warrant incorporation of them into comprehensive clinical genetic testing strategies.

Lastly, to further augment the search space for germline inherited risk events, we systematically characterized rare germline CNVs in RCC patients. Rare germline CNVs are known to increase susceptibility for different cancers^84-89^ and a few RCC studies reported CNVs detected in *FLCN* and *VHL* in RCC patients^90,91^. However, a systematic characterization of germline CNVs using WES data has not been fully explored in RCC despite the wider availability of whole exome sequencing data and improved methods for germline CNV detection. Here, we successfully identified 18 rare germline copy number duplications and deletions in CPGs from the whole-exome sequenced samples, including 4 CNVs in RCC risk genes *FH, SDHA*, and *VHL*. With the widespread use of WES and improvement in CNV identification methods, investigation of germline SNVs and short insertions and deletions together with cryptic splice and CNVs should be considered as a routine testing strategy for RCC inherited risk assessment and possibly across cancer types.

## LIMITATIONS

The current study has several limitations. First, the findings from our gene-burden analyses merit validation in additional independent case and control cohorts, particularly in larger and more diverse patient populations. Indeed, even for the sub-European ancestry identification, we did not have the means to distinguish Northwestern Europeans from Eastern or central Europeans who might have clustered together with the Northwestern Europeans in the 1000 Genomes Project-based inference, let alone for the myriad subpopulations on other non-European continents. In the future, more refined meta-analyses might take advantage of reference panels representing diverse ancestry groups to better address such subtle sub-continental differences. In addition, we had to constrain the analysis to 143 CPGs instead of a whole-exome-wide analysis due to the limited study power, further emphasizing the need for larger and more diverse patient cohorts. Finally, further studies comparing chromophobe RCC as distinct from papillary RCC as well as studies of other rare subtypes such as medullary or collecting duct RCCs are needed to better characterize inherited risk events in heterogeneous nccRCC subtypes.

## Data Availability

All computation tools and packages in this study are publicly available. The docker image containing all GATK tools is available at (https://hub.docker.com/r/broadinstitute/gatk/). The docker image containing the germline variant detection tool, DeepVariant can be found at (https://hub.docker.com/r/google/deepvariant). Tools and detailed usage for SpliceAI (https://github.com/Illumina/SpliceAI) and GATK-gCNV (https://github.com/theisaacwong/talkowski/tree/master/gCNV) can be found on the respective GitHub pages. All raw sequencing data for TCGA studies can be accessed with controlled access on the GDC data portal (https://portal.gdc.cancer.gov/) with approval. All raw sequencing data for the ICGC study can be accessed with controlled access on the ICGC data portal (https://dcc.icgc.org/) with approval. All raw sequencing data for CHECKMATE clinical studies and Genentech study can be downloaded from European Genome Phenome Archive (Dataset ID: EGAD00001001023) with approval. All raw sequencing data for cancer free control samples can be accessed on dbGAP Autism Sequencing Consortium (ASC) (dbGAP:phs000298.v4.p3), Framingham Cohort (dbGAP:phs000007.v32.p1), MESA Cohort (dbGAP: phs000209.v13.p3), NHLBI GO ESP: Lung Cohorts Exome Sequencing Project (dbGAP: phs000291.v2.p1). In house exome data for controls is available upon request.

## CONCLUSIONS AND RELEVANCE

This systematic population stratification-aware analysis supports the link between several RCC risk genes and elevated risk and describes distinct patterns of inherited germline and somatic variants in different RCC subtypes. Our results also call for caution when assessing the risk conferred by germline PVs in *CHEK2*. Finally, it broadens the definition of the RCC germline landscape of pathogenicity to incorporate previously underutilized germline variations.

## DATA AND CODE AVAILABILITY

All computation tools and packages in this study are publicly available. The docker image containing all GATK tools is available at (https://hub.docker.com/r/broadinstitute/gatk/). The docker image containing the germline variant detection tool, DeepVariant can be found at (https://hub.docker.com/r/google/deepvariant). Tools and detailed usage for SpliceAI (https://github.com/theisaacwong/talkowski/tree/master/gCNV) can be found on the respective GitHub pages. All raw sequencing data for TCGA studies can be accessed with controlled access on the GDC data portal (https://portal.gdc.cancer.gov/) with approval. All raw sequencing data for the ICGC study can be accessed with controlled access on the ICGC data portal (https://dcc.icgc.org/) with approval. All raw sequencing data for CHECKMATE clinical studies and Genentech study can be downloaded from European Genome-Phenome Archive (Dataset ID: EGAD00001001023) with approval. All raw sequencing data for cancer-free control samples can be accessed on dbGAP– Autism Sequencing Consortium (ASC) (dbGAP:phs000298.v4.p3), Framingham Cohort (dbGAP:phs000007.v32.p1), MESA Cohort (dbGAP: phs000209.v13.p3), NHLBI GO-ESP: Lung Cohorts Exome Sequencing Project (dbGAP: phs000291.v2.p1). In-house exome data for controls is available upon request.

## ACKNOWLEDGMENTS

We would like to thank all individuals who participated in the multiple studies from which we collected our raw sequencing data. We also would like to thank Isaac Wong and Jack Fu in Talkowski Lab for helping us trouble-shooting the GATK-gCNV. Finally, we would like to thank Dr. Alexander (Sasha) Gusev for his input on the Jewish Inference analysis using SNPWEIGHTs. This work was supported by The National Institutes of Health R37CA222574 (E.M.V), R01CA227388 (E.M.V), R50CA265182 (J.P.), Mark Foundation Emerging Leader Award, the Department of Defense Physician Research Award (W81XWH-21-1-0084, PC200150) (S.H.A), and the Department of Defense Idea Development Award - Early-Career Investigator (KC210042/W81XWH-22-1-0455) (S.H.A), Alex’s Lemonade Stand Foundation Young Investigator Grant (R.G.), and the Wong Family Award in Translational Oncology (R.G.). The funding organizations were not responsible for the design and conduct of the study; collection, management, analysis, and interpretation of the data; preparation, review, or approval of the manuscript; and decision to submit the manuscript for publication.

## AUTHORS’ CONTRIBUTION

[Z. ElBakouny, B. Reardon, N. Moore, C. Labaki, D. Braun, T. K. Choueiri] are responsible for the acquisition of sequencing and clinical data and the enrollment of patients. [S. Han, S. Camp, S. H. AlDubayan, H. Chu, N. Moore, E. Kofman, B. Reardon] created the computational pipeline and processed the raw genetic data. [S. Han, S. H. AlDubayan] performed all analyses and interpretations of data. [S. Han, S. H. AlDubayan, E. Van Allen] drafted the manuscript. [S. Han] prepared the figures. [C. Ricker, J. Park] generated somatic variant data for CHECKMATE samples. [R. Gillani, R. Collins, H. Chu] provided advice and discussion imperative for the intellectual content of the manuscript. All authors reviewed and edited the manuscript.

## ETHICS DECLARATION

All individuals in this study consented to institutional review board-approved protocol (IRB# 20-293) that allowed for comprehensive genetic analysis of germline samples (Methods). This study conforms to the Declaration of Helsinki.

---

**Supplementary Figure 1.**
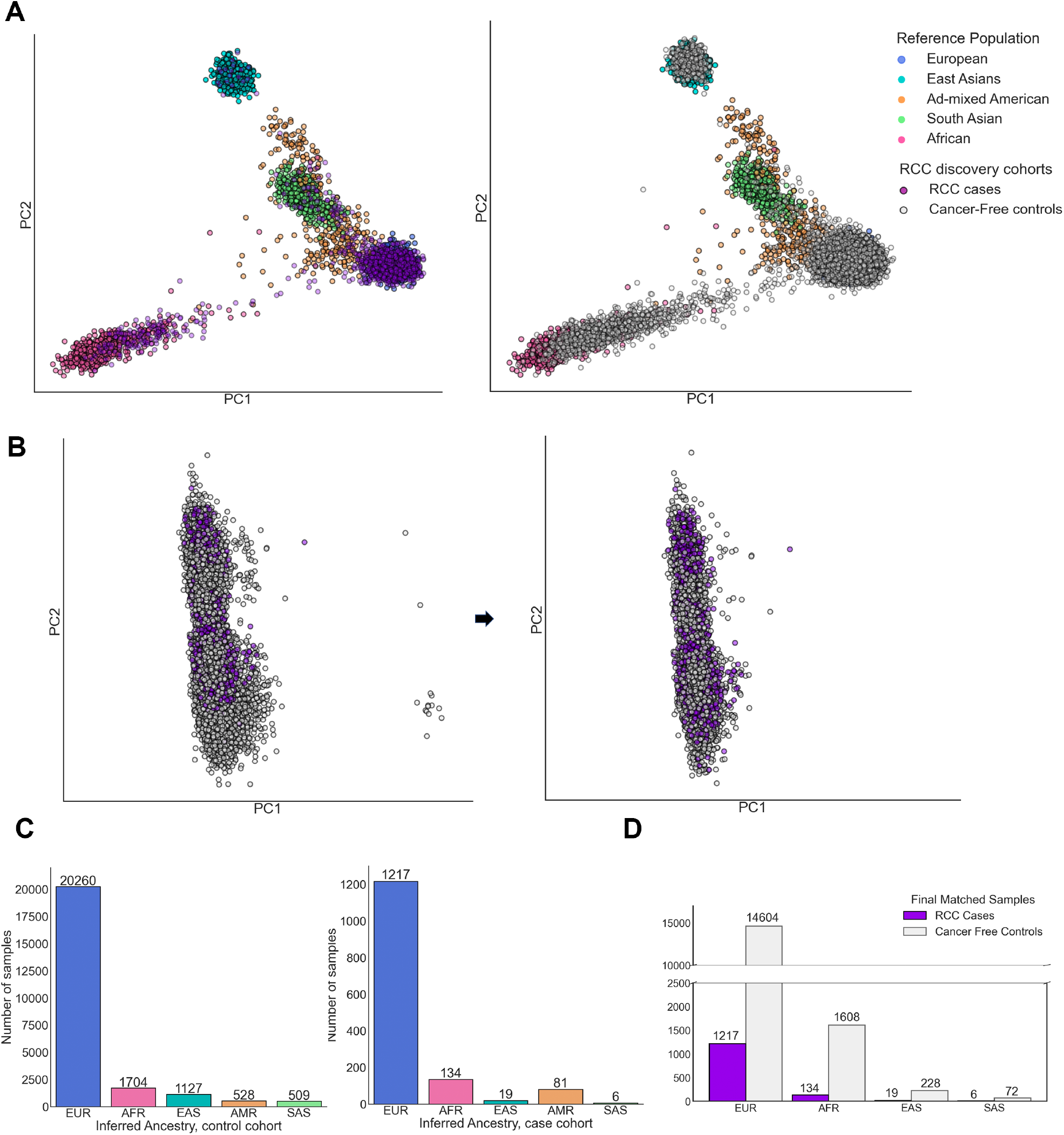
Genetic ancestry inference and ancestry matching of cases and controls. All available RCC patients were ancestry-matched with cancer-free controls, and patients with RCC subtypes other than clear cell, papillary, or chromophobe RCC were excluded from the final matched set of cases and controls. A)Projection of cases and controls on the first two principal components (PCs) along with five broad continental reference clusters formed using the samples of the 1000 Genomes project. B)Ancestry pair-matching of the cases and controls identified as European in the inference. Control samples closest to each case were identified using the top 10 PCs and the rest was excluded C)Continental ancestry assignment of cases and controls. Each sample was assigned to one of the five continental ancestries (EUR: European, AFR: African, EAS: East Asian, AMR: Admixed American, SAS: South Asian) after the first round of PCA and random forest. (D) Cases and controls were matched with a 1:12 ratio in each ancestry group. To secure the largest ratio possible between cases and controls, AMR cases and controls were excluded

**Supplementary Figure 2.**
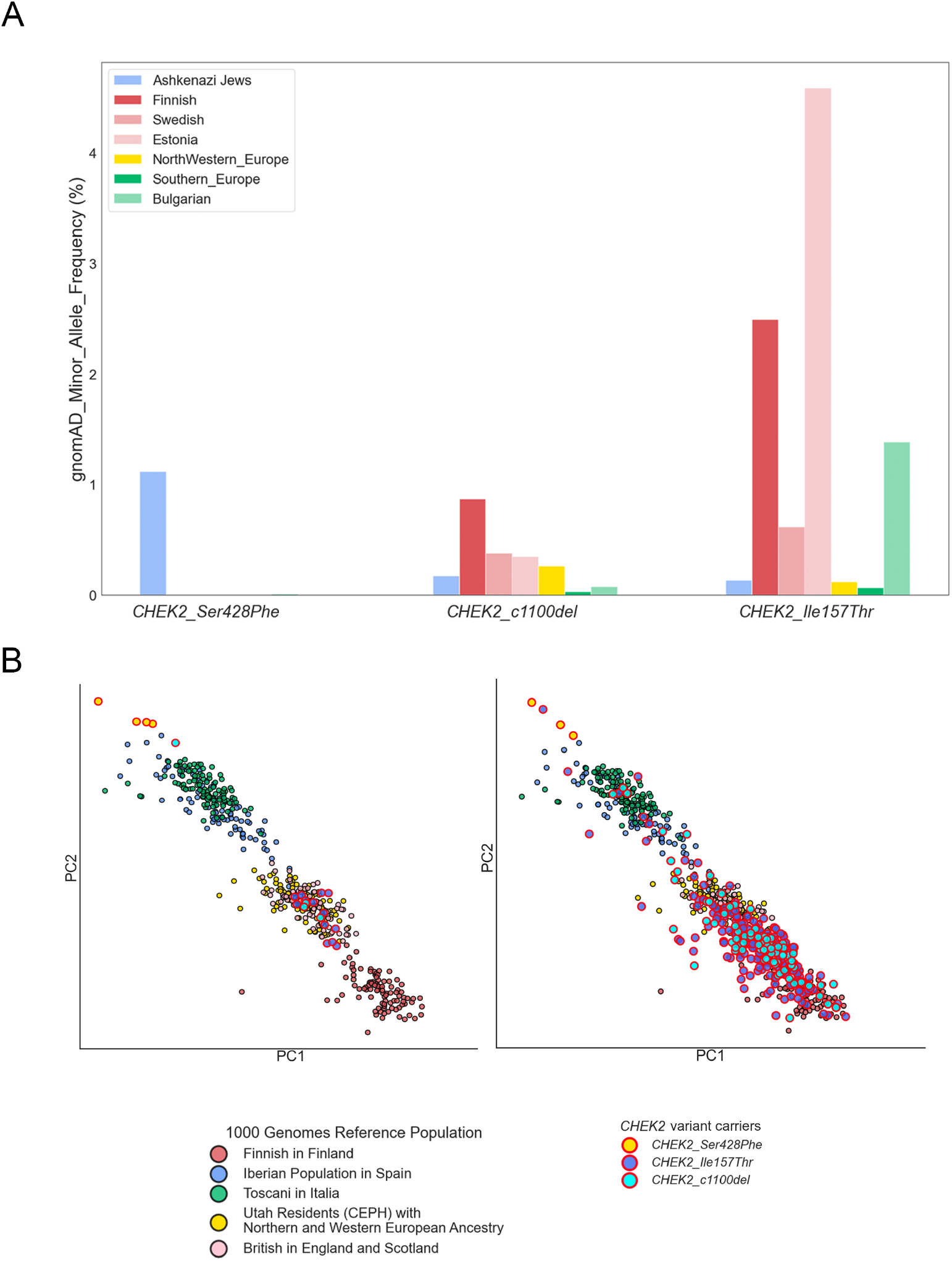
Substantial variation in population allele frequencies of *CHEK2* founder variants in different European populations. A)gnomAD minor allele frequency in % for the three *CHEK2* founder variants indicating the wide range of population frequency in different European groups B)Distribution of the three *CHEK2* founder variants in RCC cases (Left) and in cancer-free controls (Right). Reference sub-European clusters were formed using 1000 Genomes European samples with corresponding sub-European labels.

**Supplementary Figure 3.**
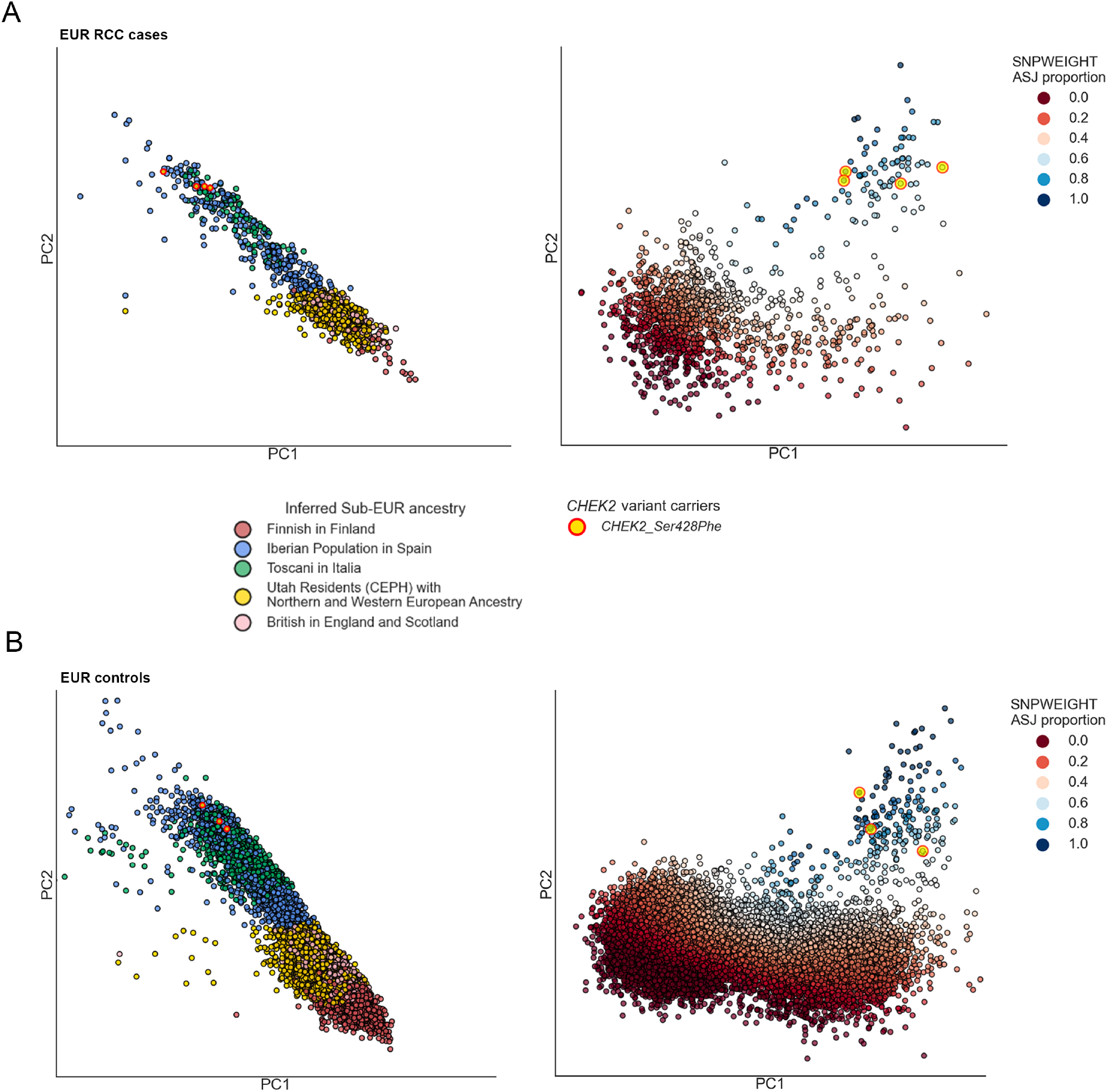
SNPWEIGHSTs sub-European ancestry inference. Ashkenazi Jewish individuals, which were unable to be identified using PCA and random forest with 1000 Genomes were identified using SNPweights. A)Left - Projection of the first two PCs for all European RCC cases color coded with inferred sub-EUR ancestry. All 4 carriers of *CHEK2* p.Ser428Phe clustered with Southern European individuals. Right - Projection of the two PCs from SNPweights with color coding indicating the level of inferred Ashkenazi Jewish proportion. All *CHEK2* p.Ser428Phe carriers fell within a cluster with a high (>0.6) ASJ proportion B)Same as A for the European controls.

**Supplementary Figure 4.**
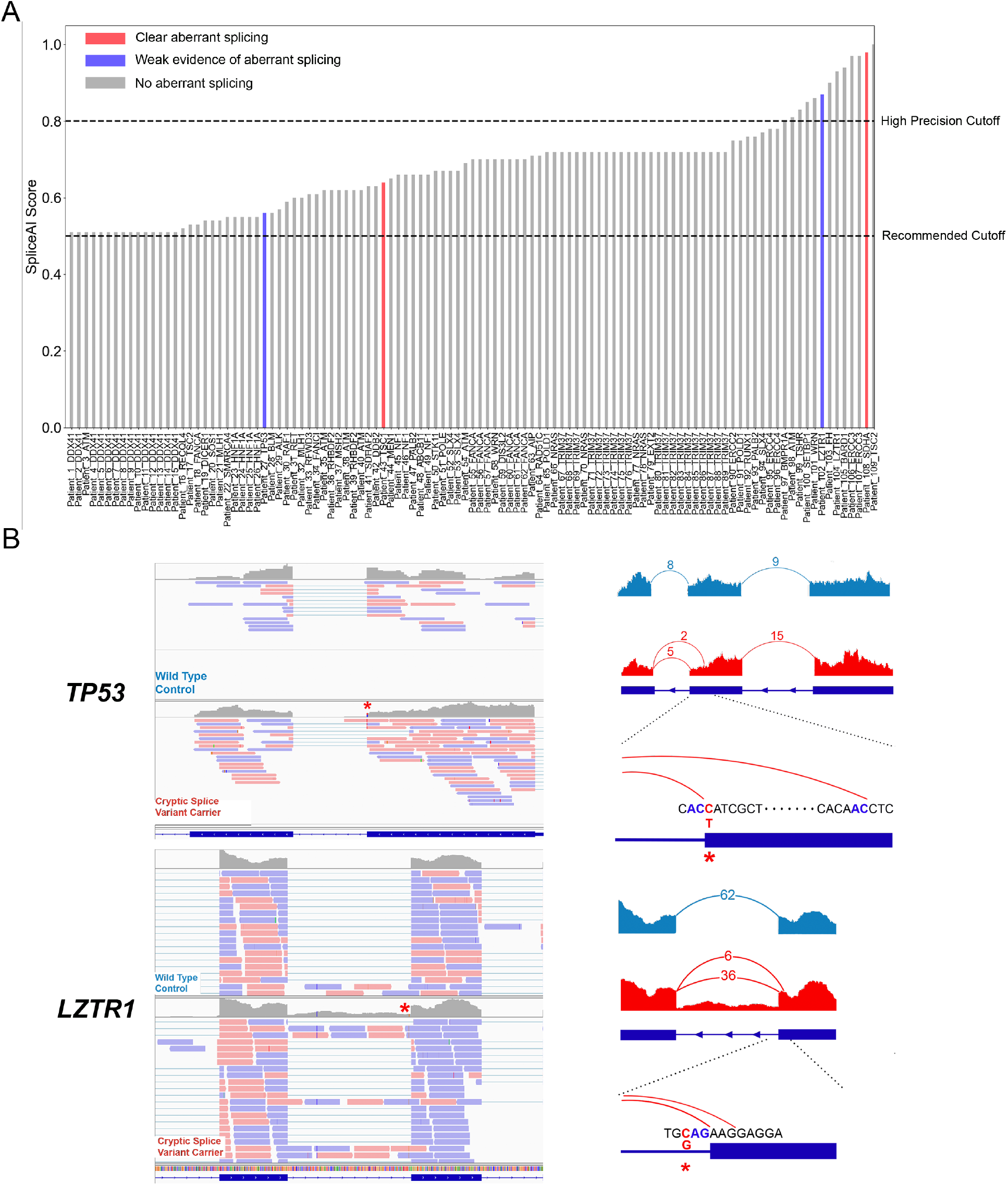
Validation of cryptic splice variants identified in the TCGA and CHECKMATE samples. A)mRNA sequencing data of 89 TCGA and CHECKMATE samples with identified cryptic splice variants were evaluated to confirm the effect of the variants. Most of the putative cryptic splice variants didn’t show evidence of aberrant splicing B)A cryptic splice variant in *TP53* and another in *LZTR1* shows aberrant splicing induced by donor loss and acceptor loss respectively, but the number of junction split reads were too low to confidently support the presence of aberrant splicing.

**Supplementary Figure 5.**
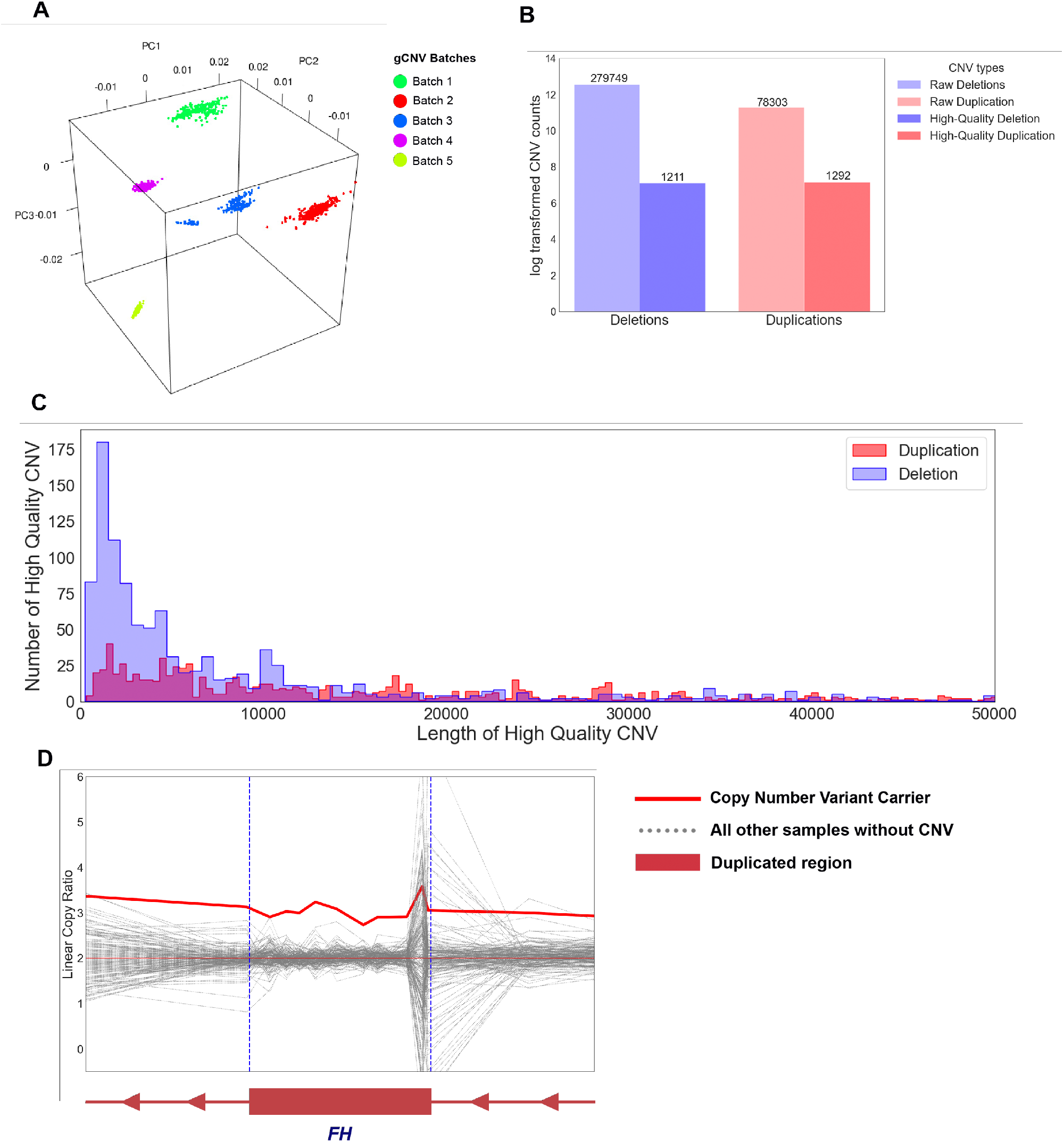
Germline copy number variant detection from whole exome sequencing data. A)PCA on sequencing baits formed 5 different clusters to run GATK-gCNV B)Bar plots indicate the number of raw deletions and duplications as well as high-quality CNVs after stringent filtering steps C)Histogram summarizing the length of high-quality duplications and deletions D)The large duplication of 56kbp covering 7 different genes including *FH*

## REFERENCES

1. Padala SA, Barsouk A, Thandra KC, et al. Epidemiology of Renal Cell Carcinoma. World J Oncol. Jun 2020;11(3):79–87. doi:10.14740/wjon1279

2. Mucci LA, Hjelmborg JB, Harris JR, et al. Familial Risk and Heritability of Cancer Among Twins in Nordic Countries. JAMA. Jan 5 2016;315(1):68–76. doi:10.1001/jama.2015.17703

3. Haas NB, Nathanson KL. Hereditary kidney cancer syndromes. Adv Chronic Kidney Dis. Jan 2014;21(1):81–90. doi:10.1053/j.ackd.2013.10.001

4. Schmidt LS, Linehan WM. Genetic predisposition to kidney cancer. Semin Oncol. Oct 2016;43(5):566–574. doi:10.1053/j.seminoncol.2016.09.001

5. Nguyen KA, Syed JS, Espenschied CR, et al. Advances in the diagnosis of hereditary kidney cancer: Initial results of a multigene panel test. Cancer. Nov 15 2017;123(22):4363–4371. doi:10.1002/cncr.30893

6. Carlo MI, Mukherjee S, Mandelker D, et al. Prevalence of Germline Mutations in Cancer Susceptibility Genes in Patients With Advanced Renal Cell Carcinoma. JAMA Oncol. Sep 1 2018;4(9):1228–1235. doi:10.1001/jamaoncol.2018.1986

7. Wu J, Wang H, Ricketts CJ, et al. Germline mutations of renal cancer predisposition genes and clinical relevance in Chinese patients with sporadic, early-onset disease. Cancer. Apr 1 2019;125(7):1060–1069. doi:10.1002/cncr.31908

8. Hartman TR, Demidova EV, Lesh RW, et al. Prevalence of pathogenic variants in DNA damage response and repair genes in patients undergoing cancer risk assessment and reporting a personal history of early-onset renal cancer. Sci Rep. Aug 11 2020;10(1):13518. doi:10.1038/s41598-020-70449-5

9. Abou Alaiwi S, Nassar AH, Adib E, et al. Trans-ethnic variation in germline variants of patients with renal cell carcinoma. Cell Rep. Mar 30 2021;34(13):108926. doi:10.1016/j.celrep.2021.108926

10. Truong H, Sheikh R, Kotecha R, et al. Germline Variants Identified in Patients with Early-onset Renal Cell Carcinoma Referred for Germline Genetic Testing. Eur Urol Oncol. Dec 2021;4(6):993–1000. doi:10.1016/j.euo.2021.09.005

11. Yngvadottir B, Andreou A, Bassaganyas L, et al. Frequency of pathogenic germline variants in cancer susceptibility genes in 1336 renal cell carcinoma cases. Hum Mol Genet. Aug 25 2022;31(17):3001–3011. doi:10.1093/hmg/ddac089

12. Bychkovsky BL, Agaoglu NB, Horton C, et al. Differences in Cancer Phenotypes Among Frequent CHEK2 Variants and Implications for Clinical Care-Checking CHEK2. JAMA Oncol. Sep 22 2022;doi:10.1001/jamaoncol.2022.4071

13. Sato Y, Yoshizato T, Shiraishi Y, et al. Integrated molecular analysis of clear-cell renal cell carcinoma. Nat Genet. Aug 2013;45(8):860–7. doi:10.1038/ng.2699

14. Motzer RJ, Jonasch E, Boyle S, et al. NCCN Guidelines Insights: Kidney Cancer, Version 1.2021. J Natl Compr Canc Netw. Sep 2020;18(9):1160–1170. doi:10.6004/jnccn.2020.0043

15. Persyn E, Redon R, Bellanger L, Dina C. The impact of a fine-scale population stratification on rare variant association test results. PLoS One. 2018;13(12):e0207677. doi:10.1371/journal.pone.0207677

16. Cancer Genome Atlas Research N. Comprehensive molecular characterization of clear cell renal cell carcinoma. Nature. Jul 4 2013;499(7456):43–9. doi:10.1038/nature12222

17. Cancer Genome Atlas Research N, Linehan WM, Spellman PT, et al. Comprehensive Molecular Characterization of Papillary Renal-Cell Carcinoma. N Engl J Med. Jan 14 2016;374(2):135–45. doi:10.1056/NEJMoa1505917

18. Davis CF, Ricketts CJ, Wang M, et al. The somatic genomic landscape of chromophobe renal cell carcinoma. Cancer Cell. Sep 8 2014;26(3):319–330. doi:10.1016/j.ccr.2014.07.014

19. Motzer RJ, Escudier B, McDermott DF, et al. Nivolumab versus Everolimus in Advanced Renal-Cell Carcinoma. N Engl J Med. Nov 5 2015;373(19):1803–13. doi:10.1056/NEJMoa1510665

20. Motzer RJ, Rini BI, McDermott DF, et al. Nivolumab for Metastatic Renal Cell Carcinoma: Results of a Randomized Phase II Trial. J Clin Oncol. May 1 2015;33(13):1430–7. doi:10.1200/JCO.2014.59.0703

21. Choueiri TK, Fishman MN, Escudier B, et al. Immunomodulatory Activity of Nivolumab in Metastatic Renal Cell Carcinoma. Clin Cancer Res. Nov 15 2016;22(22):5461–5471. doi:10.1158/1078-0432.CCR-15-2839

22. Consortium ITP-CAoWG. Pan-cancer analysis of whole genomes. Nature. Feb 2020;578(7793):82–93. doi:10.1038/s41586-020-1969-6

23. Durinck S, Stawiski EW, Pavia-Jimenez A, et al. Spectrum of diverse genomic alterations define non-clear cell renal carcinoma subtypes. Nat Genet. Jan 2015;47(1):13–21. doi:10.1038/ng.3146

24. Genomes Project C, Auton A, Brooks LD, et al. A global reference for human genetic variation. Nature. Oct 1 2015;526(7571):68–74. doi:10.1038/nature15393

25. McKenna A, Hanna M, Banks E, et al. The Genome Analysis Toolkit: a MapReduce framework for analyzing next-generation DNA sequencing data. Genome Res. Sep 2010;20(9):1297–303. doi:10.1101/gr.107524.110

26. Poplin R, Chang PC, Alexander D, et al. A universal SNP and small-indel variant caller using deep neural networks. Nat Biotechnol. Nov 2018;36(10):983–987. doi:10.1038/nbt.4235

27. AlDubayan SH, Conway JR, Camp SY, et al. Detection of Pathogenic Variants With Germline Genetic Testing Using Deep Learning vs Standard Methods in Patients With Prostate Cancer and Melanoma. JAMA. Nov 17 2020;324(19):1957–1969. doi:10.1001/jama.2020.20457

28. Camp SY, Kofman E, Reardon B, et al. Evaluating the molecular diagnostic yield of joint genotyping-based approach for detecting rare germline pathogenic and putative loss-of-function variants. Genet Med. May 2021;23(5):918–926. doi:10.1038/s41436-020-01074-w

29. Conomos MP, Miller MB, Thornton TA. Robust inference of population structure for ancestry prediction and correction of stratification in the presence of relatedness. Genet Epidemiol. May 2015;39(4):276–93. doi:10.1002/gepi.21896

30. Conomos MP, Reiner AP, Weir BS, Thornton TA. Model-free Estimation of Recent Genetic Relatedness. Am J Hum Genet. Jan 7 2016;98(1):127–48. doi:10.1016/j.ajhg.2015.11.022

31. Team H. Hail 0.2. 2021:https://github.com/hail-is/hail.

32. Chen CY, Pollack S, Hunter DJ, Hirschhorn JN, Kraft P, Price AL. Improved ancestry inference using weights from external reference panels. Bioinformatics. Jun 1 2013;29(11):1399–406. doi:10.1093/bioinformatics/btt144

33. McLaren W, Gil L, Hunt SE, et al. The Ensembl Variant Effect Predictor. Genome Biol. Jun 6 2016;17(1):122. doi:10.1186/s13059-016-0974-4

34. Liu X, Li C, Mou C, Dong Y, Tu Y. dbNSFP v4: a comprehensive database of transcript-specific functional predictions and annotations for human nonsynonymous and splice-site SNVs. Genome Med. Dec 2 2020;12(1):103. doi:10.1186/s13073-020-00803-9

35. Landrum MJ, Lee JM, Benson M, et al. ClinVar: improving access to variant interpretations and supporting evidence. Nucleic Acids Res. Jan 4 2018;46(D1):D1062–D1067. doi:10.1093/nar/gkx1153

36. Richards S, Aziz N, Bale S, et al. Standards and guidelines for the interpretation of sequence variants: a joint consensus recommendation of the American College of Medical Genetics and Genomics and the Association for Molecular Pathology. Genet Med. May 2015;17(5):405–24. doi:10.1038/gim.2015.30

37. Kopanos C, Tsiolkas V, Kouris A, et al. VarSome: the human genomic variant search engine. Bioinformatics. Jun 1 2019;35(11):1978–1980. doi:10.1093/bioinformatics/bty897

38. Perktold J S, Skipper. statsmodels: Econometric and statistical modeling with python. presented at: Proceedings of the 9th Python in Science Conference; 2010;

39. Virtanen P, Gommers R, Oliphant TE, et al. SciPy 1.0: fundamental algorithms for scientific computing in Python. Nat Methods. Mar 2020;17(3):261–272. doi:10.1038/s41592-019-0686-2

40. Viechtbauer W. Conducting Meta-Analyses in R with the metafor Package. 2010;

41. Crowdis J, He MX, Reardon B, Van Allen EM. CoMut: visualizing integrated molecular information with comutation plots. Bioinformatics. Aug 1 2020;36(15):4348–4349. doi:10.1093/bioinformatics/btaa554

42. Cibulskis K, Lawrence MS, Carter SL, et al. Sensitive detection of somatic point mutations in impure and heterogeneous cancer samples. Nat Biotechnol. Mar 2013;31(3):213–9. doi:10.1038/nbt.2514

43. Cibulskis K, McKenna A, Fennell T, Banks E, DePristo M, Getz G. ContEst: estimating cross-contamination of human samples in next-generation sequencing data. Bioinformatics. Sep 15 2011;27(18):2601–2. doi:10.1093/bioinformatics/btr446

44. Saunders CT, Wong WS, Swamy S, Becq J, Murray LJ, Cheetham RK. Strelka: accurate somatic small-variant calling from sequenced tumor-normal sample pairs. Bioinformatics. Jul 15 2012;28(14):1811–7. doi:10.1093/bioinformatics/bts271

45. Costello M, Pugh TJ, Fennell TJ, et al. Discovery and characterization of artifactual mutations in deep coverage targeted capture sequencing data due to oxidative DNA damage during sample preparation. Nucleic Acids Res. Apr 1 2013;41(6):e67. doi:10.1093/nar/gks1443

46. Taylor-Weiner A, Stewart C, Giordano T, et al. DeTiN: overcoming tumor-in-normal contamination. Nat Methods. Jul 2018;15(7):531–534. doi:10.1038/s41592-018-0036-9

47. Landau DA, Carter SL, Stojanov P, et al. Evolution and impact of subclonal s in chronic lymphocytic leukemia. Cell. Feb 14 2013;152(4):714–26. doi:10.1016/j.cell.2013.01.019

48. Lawrence MS, Stojanov P, Mermel CH, et al. Discovery and saturation analysis of cancer genes across 21 tumour types. Nature. Jan 23 2014;505(7484):495–501. doi:10.1038/nature12912

49. Carter SL, Cibulskis K, Helman E, et al. Absolute quantification of somatic DNA alterations in human cancer. Nat Biotechnol. May 2012;30(5):413–21. doi:10.1038/nbt.2203

50. Ramos AH, Lichtenstein L, Gupta M, et al. Oncotator: cancer variant annotation tool. Hum Mutat. Apr 2015;36(4):E2423–9. doi:10.1002/humu.22771

51. Braun DA, Street K, Burke KP, et al. Progressive immune dysfunction with advancing disease stage in renal cell carcinoma. Cancer Cell. May 10 2021;39(5):632–648 e8. doi:10.1016/j.ccell.2021.02.013

52. Shen R, Seshan VE. FACETS: allele-specific copy number and clonal heterogeneity analysis tool for high-throughput DNA sequencing. Nucleic Acids Res. Sep 19 2016;44(16):e131. doi:10.1093/nar/gkw520

53. Liu D, Schilling B, Liu D, et al. Integrative molecular and clinical modeling of clinical outcomes to PD1 blockade in patients with metastatic melanoma. Nat Med. Dec 2019;25(12):1916–1927. doi:10.1038/s41591-019-0654-5

54. Jaganathan K, Kyriazopoulou Panagiotopoulou S, McRae JF, et al. Predicting Splicing from Primary Sequence with Deep Learning. Cell. Jan 24 2019;176(3):535–548 e24. doi:10.1016/j.cell.2018.12.015

55. Babadi M, Fu JM, Lee SK, et al. GATK-gCNV: A Rare Copy Number Variant Discovery Algorithm and Its Application to Exome Sequencing in the UK Biobank. bioRxiv. 2022;doi:10.1101/2022.08.25.504851

56. Karczewski KJ, Francioli LC, Tiao G, et al. The mutational constraint spectrum quantified from variation in 141,456 humans. Nature. May 2020;581(7809):434–443. doi:10.1038/s41586-020-2308-7

57. Jonasch E, Walker CL, Rathmell WK. Clear cell renal cell carcinoma ontogeny and mechanisms of lethality. Nat Rev Nephrol. Apr 2021;17(4):245–261. doi:10.1038/s41581-020-00359-2

58. Melhem-Bertrandt A, Bojadzieva J, Ready KJ, et al. Early onset HER2-positive breast cancer is associated with germline TP53 mutations. Cancer. Feb 15 2012;118(4):908–13. doi:10.1002/cncr.26377

59. Pearlman R, Frankel WL, Swanson B, et al. Prevalence and Spectrum of Germline Cancer Susceptibility Gene Mutations Among Patients With Early-Onset Colorectal Cancer. JAMA Oncol. Apr 1 2017;3(4):464–471. doi:10.1001/jamaoncol.2016.5194

60. Liu YL, Cadoo KA, Maio A, et al. Early age of onset and broad cancer spectrum persist in MSH6- and PMS2-associated Lynch syndrome. Genet Med. Jun 2022;24(6):1187–1195. doi:10.1016/j.gim.2022.02.016

61. Reckamp KL, Behrendt CE, Slavin TP, et al. Germline mutations and age at onset of lung adenocarcinoma. Cancer. Aug 1 2021;127(15):2801–2806. doi:10.1002/cncr.33573

62. ZK Stadler AM, A Padunan, et al. Germline mutation prevalence in young adults with cancer. Presented at: American Association for Cancer Research Virtual Annual Meeting II. 2020;

63. Weitzel JN, Blazer KR, MacDonald DJ, Culver JO, Offit K. Genetics, genomics, and cancer risk assessment: State of the Art and Future Directions in the Era of Personalized Medicine. CA Cancer J Clin. Sep-Oct 2011;61(5):327–59. doi:10.3322/caac.20128

64. Collins RL, Brand H, Karczewski KJ, et al. A structural variation reference for medical and population genetics. Nature. May 2020;581(7809):444–451. doi:10.1038/s41586-020-2287-8

65. Ricketts CJ, Crooks DR, Sourbier C, Schmidt LS, Srinivasan R, Linehan WM. SnapShot: Renal Cell Carcinoma. Cancer Cell. Apr 11 2016;29(4):610–610 e1. doi:10.1016/j.ccell.2016.03.021

66. Karczewski KJ, Weisburd B, Thomas B, et al. The ExAC browser: displaying reference data information from over 60 000 exomes. Nucleic Acids Res. Jan 4 2017;45(D1):D840–D845. doi:10.1093/nar/gkw971

67. Tintle N, Aschard H, Hu I, Nock N, Wang H, Pugh E. Inflated type I error rates when using aggregation methods to analyze rare variants in the 1000 Genomes Project exon sequencing data in unrelated individuals: summary results from Group 7 at Genetic Analysis Workshop 17. Genet Epidemiol. 2011;35 Suppl 1(Suppl 1):S56–60. doi:10.1002/gepi.20650

68. Mathieson I, McVean G. Differential confounding of rare and common variants in spatially structured populations. Nat Genet. Feb 5 2012;44(3):243–6. doi:10.1038/ng.1074

69. Jiang Y, Epstein MP, Conneely KN. Assessing the impact of population stratification on association studies of rare variation. Hum Hered. 2013;76(1):28–35. doi:10.1159/000353270

70. Zhang Y, Guan W, Pan W. Adjustment for population stratification via principal components in association analysis of rare variants. Genet Epidemiol. Jan 2013;37(1):99–109. doi:10.1002/gepi.21691

71. Consortium CBCC-C. CHEK2*1100delC and susceptibility to breast cancer: a collaborative analysis involving 10,860 breast cancer cases and 9,065 controls from 10 studies. Am J Hum Genet. Jun 2004;74(6):1175–82. doi:10.1086/421251

72. Kenneth Offit HP, Tomas Kirchhoff, Prema Kolachana, Beth Rapaport, Peter Gregersen, Steven Johnson, Orit Yossepowitch, Helen Huang Jaya Satagopan, Mark Robson, Lauren Scheuer, Khedoudja Nafa and Nathan Ellis. Frequency of CHEK2*1100delC in New York breast cancer cases and controls. BMC Medical Genetics. 2003;

73. Mateus Pereira LH, Sigurdson AJ, Doody MM, et al. CHEK2:1100delC and female breast cancer in the United States. Int J Cancer. Nov 10 2004;112(3):541–3. doi:10.1002/ijc.20439

74. Neuhausen S, Dunning A, Steele L, et al. Role of CHEK2*1100delC in unselected series of non-BRCA1/2 male breast cancers. Int J Cancer. Jan 20 2004;108(3):477–8. doi:10.1002/ijc.11385

75. Osorio A, Rodriguez-Lopez R, Diez O, et al. The breast cancer low-penetrance allele 1100delC in the CHEK2 gene is not present in Spanish familial breast cancer population. Int J Cancer. Jan 1 2004;108(1):54–6. doi:10.1002/ijc.11414

76. Vahteristo P, Bartkova J, Eerola H, et al. A CHEK2 genetic variant contributing to a substantial fraction of familial breast cancer. Am J Hum Genet. Aug 2002;71(2):432–8. doi:10.1086/341943

77. Yael Laitman BK, Ephrat Levy Lahad, Moshe Z. Papa and Eitan Friedman. Germline CHEK2 Mutations in Jewish Ashkenazi Women at High Risk for Breast Cancer. Israel Medical Association Journal. 2007;

78. Stolarova L, Kleiblova P, Janatova M, et al. CHEK2 Germline Variants in Cancer Predisposition: Stalemate Rather than Checkmate. Cells. Dec 12 2020;9(12)doi:10.3390/cells9122675

79. Margolin S, Eiberg H, Lindblom A, Bisgaard ML. CHEK2 1100delC is prevalent in Swedish early onset familial breast cancer. BMC Cancer. Aug 17 2007;7:163. doi:10.1186/1471-2407-7-163

80. Rashid MU, Jakubowska A, Justenhoven C, et al. German populations with infrequent CHEK2*1100delC and minor associations with early-onset and familial breast cancer. Eur J Cancer. Dec 2005;41(18):2896–903. doi:10.1016/j.ejca.2005.04.049

81. Rogier A. Oldenburg KK-J, Jaennelle Kraan,Hans Morreau, Jan G. M. Klijn, Nicoline Hoogerbrugge, Marjolein J. L. Ligtenberg, Christi J. van Asperen, Hans F. A. Vasen, Carel Meijers,, Hanne Meijers-Heijboer THdB, Cees J. Cornelisse, and Peter Devilee. The CHEK2*1100delC Variant Acts as a Breast Cancer Risk Modifier in Non-BRCA1/BRCA2 Multiple-Case Families. CANCER RESEARCH. 2003;

82. Lee M, Roos P, Sharma N, et al. Systematic Computational Identification of Variants That Activate Exonic and Intronic Cryptic Splice Sites. Am J Hum Genet. May 4 2017;100(5):751–765. doi:10.1016/j.ajhg.2017.04.001

83. Cooper TA, Wan L, Dreyfuss G. RNA and disease. Cell. Feb 20 2009;136(4):777–93. doi:10.1016/j.cell.2009.02.011

84. Walker LC, Pearson JF, Wiggins GA, Giles GG, Hopper JL, Southey MC. Increased genomic burden of germline copy number variants is associated with early onset breast cancer: Australian breast cancer family registry. Breast Cancer Res. Mar 16 2017;19(1):30. doi:10.1186/s13058-017-0825-6

85. Laitinen VH, Akinrinade O, Rantapero T, Tammela TL, Wahlfors T, Schleutker J. Germline copy number variation analysis in Finnish families with hereditary prostate cancer. Prostate. Feb 15 2016;76(3):316–24. doi:10.1002/pros.23123

86. Yoshihara K, Tajima A, Adachi S, et al. Germline copy number variations in BRCA1-associated ovarian cancer patients. Genes Chromosomes Cancer. Mar 2011;50(3):167–77. doi:10.1002/gcc.20841

87. Brea-Fernandez AJ, Fernandez-Rozadilla C, Alvarez-Barona M, et al. Candidate predisposing germline copy number variants in early onset colorectal cancer patients. Clin Transl Oncol. May 2017;19(5):625–632. doi:10.1007/s12094-016-1576-z

88. Shi J, Zhou W, Zhu B, et al. Rare Germline Copy Number Variations and Disease Susceptibility in Familial Melanoma. J Invest Dermatol. Dec 2016;136(12):2436–2443. doi:10.1016/j.jid.2016.07.023

89. Park RW, Kim TM, Kasif S, Park PJ. Identification of rare germline copy number variations over-represented in five human cancer types. Mol Cancer. Feb 3 2015;14:25. doi:10.1186/s12943-015-0292-6

90. Schneider M, Dinkelborg K, Xiao X, et al. Early onset renal cell carcinoma in an adolescent girl with germline FLCN exon 5 deletion. Fam Cancer. Jan 2018;17(1):135–139. doi:10.1007/s10689-017-0008-8

91. Matsuda D, Khoo SK, Massie A, et al. Identification of copy number alterations and its association with pathological features in clear cell and papillary RCC. Cancer Lett. Dec 18 2008;272(2):260–7. doi:10.1016/j.canlet.2008.06.015

